# CB-COMIL: interpretable prediction of melanoma immunotherapy response from multiplex immunofluorescence whole-slide images

**DOI:** 10.64898/2026.11.19.26368529

**Authors:** Priyanka Rana, Tuba N Gide, Nurudeen A Adegoke, Yizhe Mao, Serigne N. Lo, Shlomo Berkovsky, Enrico Coiera, Georgina V. Long, James S Wilmott, Sidong Liu

**Affiliations:** Centre for Health Informatics, Australian Institute of Health Innovation, Macquarie University, Sydney, NSW, Australia; Melanoma Institute Australia, The University of Sydney, Sydney, NSW, Australia; Charles Perkins Centre, The University of Sydney, Sydney, NSW, Australia; Faculty of Medicine and Health, The University of Sydney, Sydney, NSW, Australia; Mater Hospital, North Sydney, Sydney, NSW, Australia; Royal North Shore Hospital, Sydney, Australia

## Abstract

Deep learning models can predict melanoma immunotherapy response but rarely explain predictions biologically. Concept bottleneck models offer interpretable predictions, but have not been applied to multiplex immunofluorescence (mIF), and whether their learned concepts reproduce tissue measurements remains unclear. We developed CB-COMIL, which routes mIF predictions through four supervised concepts (immune:tumour ratio, CD8/SOX10 balance, CD68 relative signal and PD-L1 evasion index), decomposing each prediction exactly into concept contributions. Developed on 211 slides and evaluated on 42 slides from an independent, differently acquired test cohort, CB-COMIL achieved macro-F1 0.69 versus 0.67 for an opaque comparator (difference 0.020, 95% CI *−*0.103 to 0.144). In 41 slides with complete measurements, learned concepts correlated with measured counterparts (Spearman *ρ* 0.61–0.76), yet substituting measured values reduced macro-F1 from 0.68 to 0.48. CB-COMIL makes predictions interpretable without detectable performance loss, and our evaluation shows that concepts can faithfully determine predictions without reproducing the corresponding biological measurements.

## Introduction

Melanoma is among the most aggressive skin cancers, with an estimated 331,722 new cases and 58,667 deaths worldwide in 2022, and its incidence continues to rise [1]. Immune checkpoint blockade (ICB), a form of cancer immunotherapy, has transformed melanoma treatment by enabling the immune system to recognise and eliminate cancer cells [2]. ICB is administered either as monotherapy, typically targeting programmed cell death protein 1 (PD-1) with agents such as nivolumab, or as combination therapy targeting multiple immune checkpoints, such as nivolumab with the anti-CTLA-4 antibody ipilimumab. Yet roughly 60% of patients do not respond to monotherapy, and even in combination around 50% fail to benefit, while still enduring debilitating toxicity and substantial personal and healthcare costs [3]. Identifying likely non-responders before treatment is therefore critical, both to spare patients futile toxicity and to redirect them towards alternative therapies. This need has motivated the development of computational models, among which image-based models that capture complex patterns within the tumour microenvironment (TME) are emerging as particularly promising.

Among tissue imaging modalities, multiplex immunofluorescence (mIF) provides a basis for charac-terising the tumour microenvironment by visualising multiple immune and tumour markers within the same tissue section. By simultaneously profiling multiple biomarkers, mIF enables quantification of marker co-expression and spatial cellular relationships associated with immunotherapy response, providing information beyond that available from single-marker tissue imaging [4]. Several tumour and immune features associated with checkpoint blockade response can be characterised using mIF, including immune and tumour composition, CD8 T cell infiltration, PD-L1 expression, and macrophage composition [5, 6, 7, 8]. The relationship between macrophage composition and response depends on macrophage phenotype and treatment context [8].

A model that expressed its predictions in terms of such biologically grounded features would provide a more clinically interpretable account of its decision than one expressed only through latent image features. To analyse whole-slide mIF images and capture their high-dimensional, non-linear relationships, deep learning–based computational frameworks have recently emerged [9, 10, 11]. However, despite their predictive performance, these models typically do not expose the biological factors underlying individual predictions, limiting clinical scrutiny of their decisions.

This opacity matters most where the stakes are highest. For a decision as consequential as recommending or withholding immunotherapy, predictive accuracy alone is insufficient: the prediction should be interpretable by the pathologist, and its underlying reasoning should reflect the factors that informed the model’s decision [12]. The central question therefore shifts from whether a model performs well to whether it can be trusted to support clinical decision-making. This requires evaluating AI models not only by their predictive performance, but also by whether their reasoning is transparent and biologically meaningful.

The conventional approach to interpretability relies on post-hoc explanations, whereby a black-box model is first trained and subsequently interpreted using a separate method, such as gradient- or perturbation-based attribution (e.g., Layer-wise Relevance Propagation (LRP) [13], Class Activation Mapping (CAM) [14], Local Interpretable Model-agnostic Explanations (LIME) [15], or SHapley Additive exPlanations (SHAP) [16]). These explanations are produced after training and are not constrained by the training objective, so their faithfulness, the degree to which they reflect the model’s actual computation rather than merely appearing plausible, varies and is rarely verified. For example, some saliency based attribution methods produce visually similar maps even after the model’s learned parameters are randomised [17]. Attention weights are also widely used to explain predictions, but their faithfulness varies across architectures and tasks. In a comparative evaluation, layerwise relevance propagation, integrated gradients and perturbation methods performed better than attention weights on tests of faithfulness [18].

Faithfulness alone, however, does not provide biological interpretability. Even when an attribution map faithfully identifies regions that influence a prediction, it expresses relevance primarily at the level of image regions or features rather than through explicit, biologically defined concepts. Consequently, such maps can reveal where the model relies on the image, but provide limited insight into which biological characteristics of those regions drive the decision.

These concerns have prompted growing interest, particularly in high stakes clinical applications, in models that are interpretable by construction, where the intermediate representation driving the prediction can be examined and directly intervened upon before the final decision is formed [19]. In medical imaging, such *self-explainable* models include prototype-based and concept-based approaches [20]. Prototype-based methods explain predictions by comparing regions of a new tissue image with representative patterns learned from previously observed cases, allowing users to inspect the visual similarities underlying a decision. Both approaches offer useful insight into model behaviour, though they address different questions: prototype similarity shows which learned repre-sentative patterns a region of the image resembles, whereas a concept-based account quantifies how much each predefined biological property contributed. We therefore adopt a concept-based approach, which expresses predictions through biologically defined concepts whose individual contributions can be examined and directly intervened upon.

Concept bottleneck models (CBMs) are a representative approach of this paradigm [21]. Rather than mapping an input directly to an outcome, CBMs first predict a small set of human-specified concepts and then base the final prediction on those concepts. Consequently, the concepts are not post-hoc approximations of the model’s reasoning but constitute the decision-making process itself, providing explanations that are inherently faithful to the underlying prediction.

In computational pathology, self-interpretable multi-instance learning (SI-MIL) introduces inter-pretable structure into a deep model by pairing a MIL branch with an interpretable branch that predicts linearly from hand-crafted morphometric and spatial features, with the deep branch guiding which patches the interpretable branch sees [22]. To our knowledge, however, concept bottleneck learning has not yet been applied to predicting immunotherapy response from mIF images, where routing the prediction through biologically meaningful concepts could make the decision inter-pretable in immunological terms. More fundamentally, it remains unclear whether biologically defined concepts that faithfully govern a model’s prediction also faithfully represent the biological quantities they are intended to encode. This distinction between decision faithfulness and biological fidelity is particularly important in biomedical applications, where an interpretable representation may otherwise be mistaken for a biological measurement. It also remains unknown whether imposing such a biologically grounded concept bottleneck can preserve the predictive performance and generalisation of a strong black box model.

In this work, we propose CB-COMIL (Fig. 1), a concept bottleneck extension of our previously developed framework, Channel Optimisation with Multi-Instance Learning (COMIL) [9], to predict immunotherapy response in melanoma patients from whole-slide mIF images. The model is trained on 211 whole-slide mIF images from the Melanoma Institute Australia (MIA), each comprising seven channels: five protein biomarker channels (CD8, CD16, CD68, PD-L1 and SOX10), together with a nuclear stain (DAPI) and an autofluorescence (AF) channel. We define four concepts representing different aspects of the tumour immune microenvironment: immune signal relative to tumour signal (immune:tumour ratio), cytotoxic T-cell signal relative to melanoma-associated signal (CD8/SOX10 balance), macrophage signal relative to the CD8 and CD68 compartment (CD68 relative signal), and PD-L1 signal relative to CD8 signal (PD-L1 evasion index). These concepts were specified a priori for their biological relevance to checkpoint blockade, their measurable counterparts in the mIF panel, and their ability to provide a compact representation of the tumour immune microenvironment.

**Figure 1:**
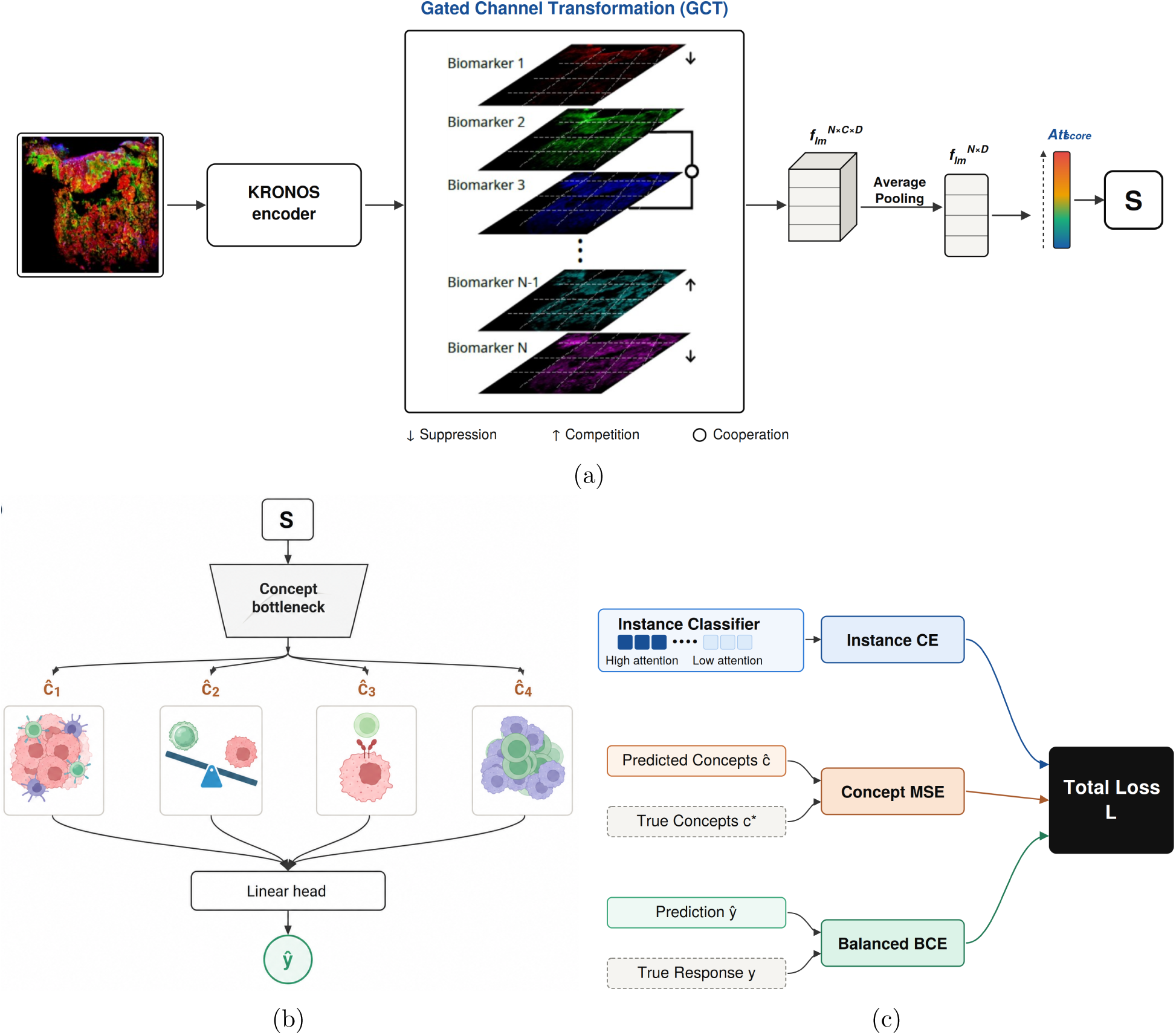
CB-COMIL, a concept bottleneck multiple-instance learning framework for immunotherapy response prediction from mIF whole-slide images. **a Marker-aware feature extraction**. Each mIF whole-slide image is encoded by KRONOS into patch-level, biomarker-resolved feature maps. A gated channel transformation (GCT) reweights the *C* biomarker channels against one another before average pooling 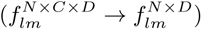; an attention head then assigns each patch a score (Att_score_) and produces the slide representation *S*. **b Concept bottleneck**. *S* is projected onto four interpretable concepts: immune–tumour ratio (*ĉ*_1_), CD8/SOX10 balance (*ĉ*_2_), CD68 relative signal (*ĉ*_3_), PD-L1 evasion index (*ĉ*_4_), which a linear head combines to give the predicted response *ŷ*. **c Training objective**. Three weighted terms are summed: instance supervision on the highest- and lowest-attention patches (cross-entropy, *µ* = 0.3), concept supervision of *ĉ* against measured concepts *c^∗^* (mean squared error, *λ* = 0.3), and response supervision of *ŷ* against the true response *y* (balanced binary cross-entropy).

CB-COMIL achieves predictive performance comparable to an opaque model while making each prediction attributable to four concepts supervised by measurable tissue quantities, and we evaluate both how much each concept contributes and how closely it corresponds to its measured counterpart. Evaluation is performed on an independent test cohort of 42 slides acquired with a different staining protocol and scanner, so the model is assessed under a measurable acquisition shift. This cohort offers a valuable opportunity for independent evaluation, particularly given the limited availability of mIF datasets with treatment response annotations and the high cost of generating such datasets. Specifically, our contributions are threefold: (1) we develop an interpretable-by-construction extension of COMIL whose macro-F1 did not differ detectably from that of the opaque model (0.69 versus 0.67; 95% CI for the difference -0.103 to +0.144); (2) we show that the model design allows each prediction logit to be decomposed exactly into concept contributions and a bias, and we use this decomposition to attribute individual predictions and quantify each concept’s influence through directional interventions; and (3) we provide empirical evidence that decision faithfulness does not necessarily imply biological fidelity: learned concept activations can govern model predictions while remaining imperfect proxies for their corresponding biological measurements, as shown by reduced predictive performance when these activations are replaced with measured values.

## Results

We evaluate CB-COMIL through four linked questions. First, does attention identify the image regions the model relies on, and can those regions alone preserve predictive performance? We test this with paired deletion and insertion analyses [23], using Integrated Gradients (IG) [24] and random rankings as benchmarks. Second, how strongly does each concept influence the prediction? We examine directional interventions, patient-level prediction decompositions and joint concept decision spaces to assess how the four learned concepts influence model decisions. Third, how closely do these concepts reflect their intended biology? We compare them with independent measurements and use corrective interventions to test whether measured values can replace learned activations. Finally, we assess whether imposing this interpretable structure preserves predictive performance on the independent test cohort, comparing CB-COMIL with the opaque backbone and feature-engineered interpretable baselines, and ablating individual components of the framework. Together, these analyses test whether interpretability by construction can be achieved without a detectable loss in performance, and what the resulting concepts do and do not represent.

Macro-F1 is the primary comparison metric throughout. It is the unweighted mean of the F1-scores for the responder and non-responder classes, giving each class equal weight regardless of its frequency. AUPRC is also reported to assess how well the model distinguishes between classes across different decision thresholds. Specificity (non-responder recall) is reported together with sensitivity (responder recall), as the model is intended to identify patients unlikely to benefit from immune checkpoint blockade.

### Attention identifies influential patches but does not establish sufficiency

We evaluated whether patches highlighted by CB-COMIL’s attention contribute to its predictions using deletion and insertion experiments [23] on the independent test cohort, comparing attention rankings with IG and a random-ranking control (Fig. 2; Supplementary Table S1).

**Figure 2:**
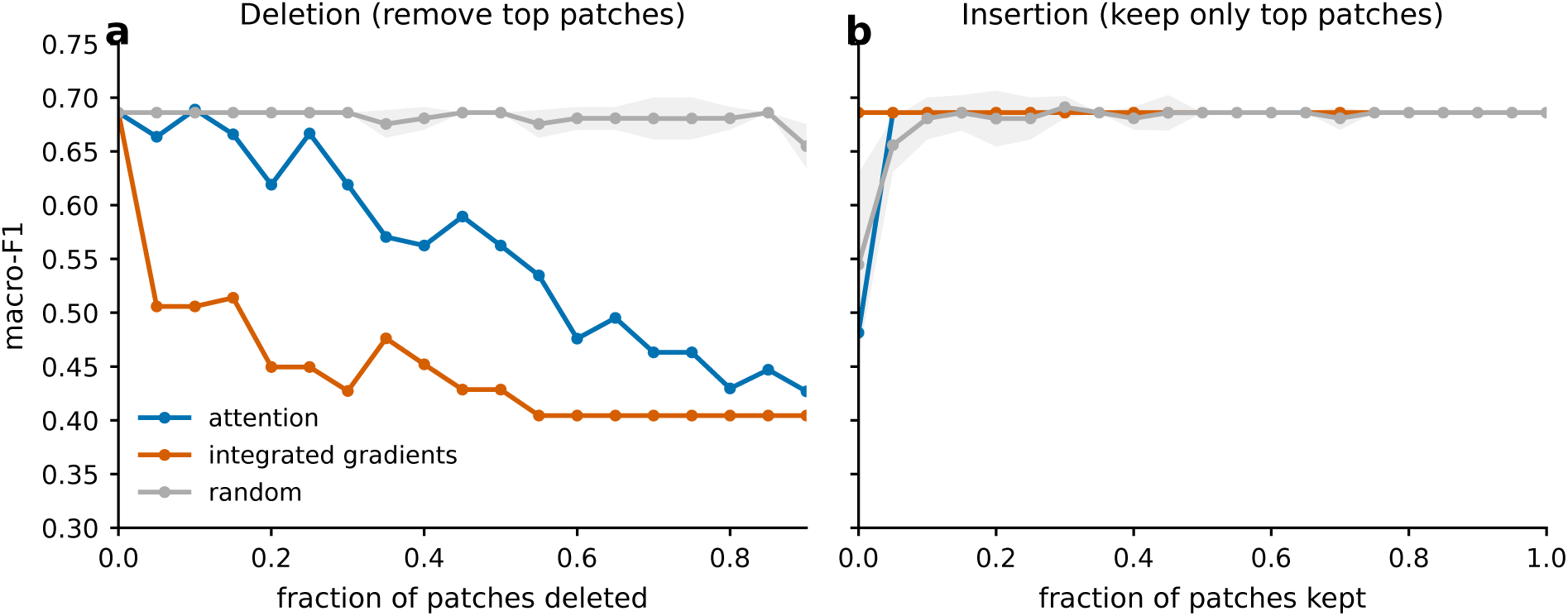
Patch-level attention identifies influential regions but does not establish sufficiency. **a,** Deletion curves obtained by progressively removing patches according to attention, Integrated Gradients (IG), or random rankings. A faster decline in predictive performance indicates greater sensitivity to removing highly ranked patches. **b,** Insertion curves obtained by progressively retaining patches according to the same rankings.

Deleting patches in descending order of attention reduced macro-F1 faster than random deletion (area under the deletion curve 0.56 versus 0.68) and changed 25.9% of predictions, compared with 0.5% for random ranking, with the same pattern for specificity, non-responder F1 and AUPRC. The largest separation was in specificity (0.51 versus 0.89), indicating that non-responder predictions were most sensitive to removal of the highest-ranked patches. IG reduced macro-F1 even faster (0.45) and changed 52.9% of predictions, with most of the decline occurring within the first 10% of patches deleted, whereas attention declined more gradually (Fig. 2a). Attention therefore identified patches that influence the prediction, but IG ranked them more effectively on most metrics.

Insertion showed no comparable advantage. Insertion macro-F1 AUCs were 0.68 for attention,

0.69 for IG and 0.68 for random ranking, and the remaining metrics differed by at most 0.01 (Supplementary Table S1). Retaining the highest-ranked patches therefore recovered performance no faster than retaining a random subset, which is consistent with predictive evidence being distributed across the slide rather than concentrated in the top-ranked patches. Patch rankings therefore indicate where the model draws its evidence, but not which biological properties of those regions drive the decision. We examine this next through the learned concepts.

### The learned concepts govern CB-COMIL’s predictions

To examine how CB-COMIL’s learned concepts contribute to its decisions, we conducted three complementary analyses: directional concept interventions to quantify their influence on predictions, patient-level decompositions to identify their individual contributions, and joint concept decision spaces to visualise how pairs of concepts shape model outputs. These analyses exploit CB-COMIL’s hard concept bottleneck, in which the response logit is computed exclusively from a linear combination of four concepts learned from image embeddings, without a prediction pathway that bypasses them. Measured mIF-derived concepts provide training supervision but are not supplied as inference inputs.

Directional interventions were performed by varying each learned concept from *−*1.5 to +1.5 standard deviations while holding the remaining concepts fixed (Table 1) to quantify the magnitude of each concept’s influence on predicted response probability. The immune–tumour ratio produced the largest change (Δ*P* = +0.40), followed by CD8/SOX10 balance (+0.22) and PD-L1 evasion (+0.17), whereas CD68 relative signal had minimal influence (+0.01). Thus, although all four concepts enter the prediction head, their influence on its output differs substantially, with the two immune-related concepts exerting the greatest influence over the evaluated intervention range.

**Table 1:** Directional concept intervention. Each concept was swept from *−*1.5 to +1.5 SD with the others held fixed; the change in mean predicted response probability P(R) quantifies each concept’s influence, and its direction follows the sign of the learned weight. The immune related concepts (immune–tumour ratio, CD8/SOX10 balance) carry the most leverage. Flips, number of the mIF samples whose prediction switched towards response.

| **Concept (learned weight)** | *P*(*R*)**: low → high** | Δ*P*(*R*) | **Flips** |
|---|---|---|---|
| Immune–tumour ratio (+0.57) | 0.29 → 0.69 | +0.40 | 41 |
| CD8/SOX10 balance (+0.33) | 0.33 → 0.55 | +0.22 | 27 |
| PD-L1 evasion index (+0.27) | 0.35 → 0.52 | +0.17 | 15 |
| CD68 relative signal (+0.02) | 0.40 → 0.41 | +0.01 | 0 |

The linear prediction head permits an exact decomposition of each patient’s response logit into a constant bias and four signed concept contributions, *θ_k_ĉ_k_*, calculated from the target-scale learned activations (Figure 3; Supplementary Table S2). Positive contributions increase the response logit, whereas negative contributions decrease it. Figure 3a illustrates these contributions in four patients representing the four confusion matrix categories. T15, a correctly classified responder (*P* (*R*) = 0.64), exhibited positive contributions from the learned immune:tumour ratio and CD8/SOX10 balance. These same concepts made substantial negative contributions in T06, a correctly classified non-responder (*P* (*R*) = 0.09), and T19, a responder incorrectly predicted as a non-responder (*P* (*R*) = 0.07). In T29, a non-responder incorrectly predicted as a responder (*P* (*R*) = 0.65), their positive contributions outweighed the negative contribution from PD-L1 evasion and combined with the constant bias to yield a responder prediction.

**Figure 3:**
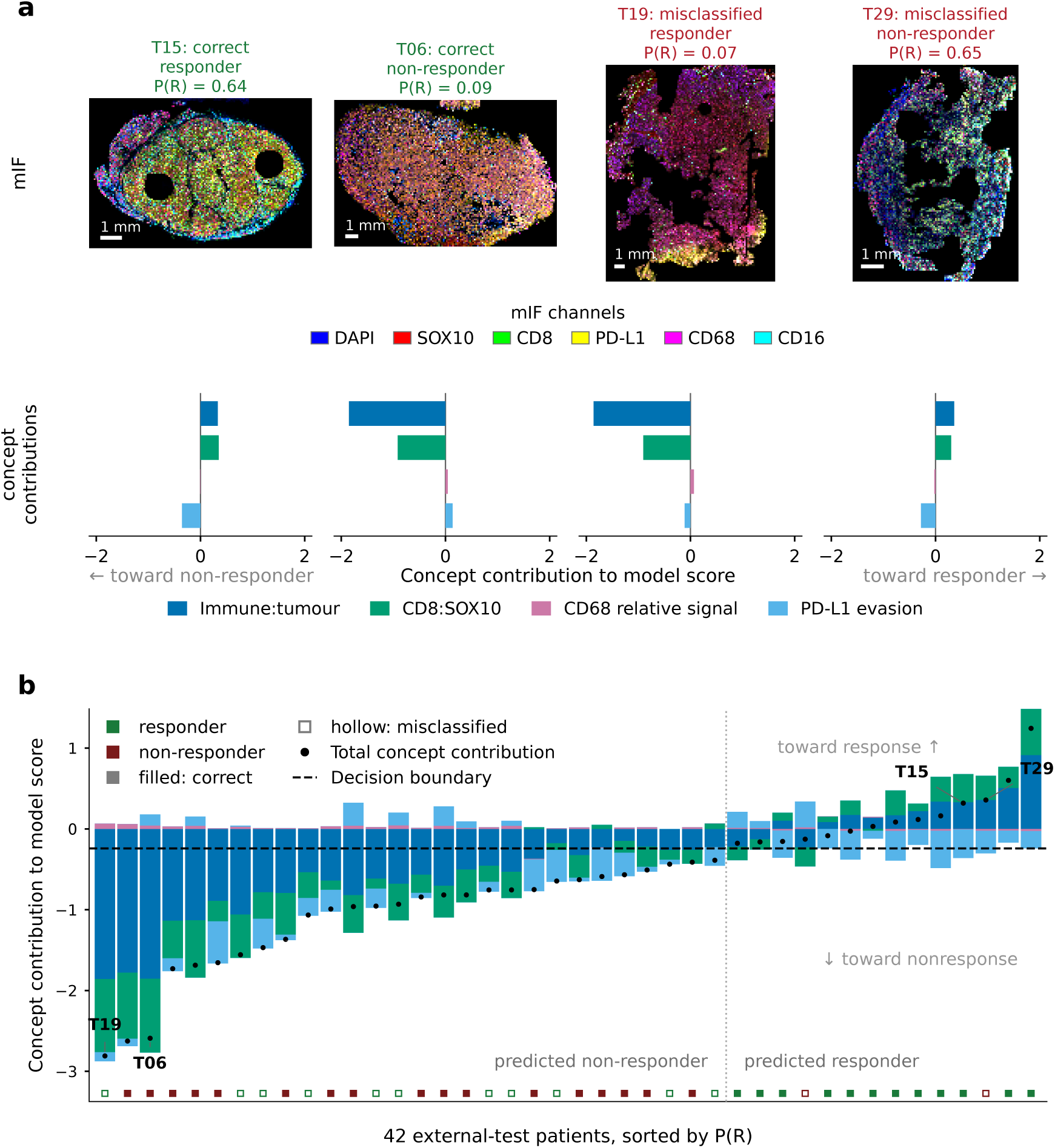
Patient-level decomposition of CB-COMIL predictions in the independent test cohort. **a**, Four cases spanning the confusion matrix: a correctly classified responder (T15), a correctly classified non-responder (T06), a missed responder (T19), and a non-responder misclassified as a responder (T29); these are illustrative, not typical, cases. The mIF composites combine DAPI (blue), SOX10 (red), CD8 (green), PD-L1 (yellow), CD68 (magenta) and CD16 (cyan). Each model logit decomposes exactly into the bias *b* and four signed concept contributions *θ_k_ĉ_k_*, computed from the target-scale learned concept activations **ĉ** received by the linear prediction head; positive contributions shift the prediction towards response and negative contributions towards non-response. CD68 relative signal forms a narrow band because its learned coefficient is near zero. **b**, Decomposition of prediction logits for all 42 patients in the independent test cohort, ordered from left to right by increasing predicted probability of response. The immune ratio and CD8 balance contribute most in absolute magnitude for most patients. Black dots show the sum of the four concept contributions before adding the intercept. The bias *b* is omitted from the bars for clarity but is included when computing *P* (R); the dashed line is therefore drawn at *−b*, where the response logit is zero and *P* (R) = 0.5, so a total contribution above this line gives a predicted responder. Panel **a** shows which concepts determine individual predictions; panel **b** shows how t_8_hose contributions position each patient relative to the decision boundary.

The cohort decomposition (Figure 3b) displays the signed concept contributions of the four learned concepts to the model score for all 42 patients in the independent test cohort, ordered by *P* (*R*). The learned immune:tumour ratio and CD8/SOX10 balance generally made the largest contributions in magnitude, whereas CD68 relative signal contributed least. Individual predictions reflected the balance of positive and negative contributions, together with the constant bias. In T29, a non-responder incorrectly predicted as a responder, positive contributions from the two immune concepts outweighed the negative PD-L1 evasion contribution. Conversely, T19 was a responder whose contributions strongly favoured non-response.

To examine the joint influence of the two concepts with the largest intervention effects and generally the largest contributions to individual predictions, we visualised the conditional decision space of the learned immune:tumour ratio and CD8/SOX10 balance (Figure 4). The background colour shows the predicted response probability, *P* (R), for each combination of these target-scale learned activations, with CD68 relative signal and PD-L1 evasion fixed at their training cohort means. Higher activations corresponded to higher predicted response probabilities, consistent with their positive contributions to the response logit. Responders generally occupied regions with higher activations, whereas non-responders tended towards lower values, although the groups overlapped. The two highlighted misclassified non-responders occupied different positions: T29 lay in a region of higher predicted response probability, whereas T33 lay close to the displayed *P* (R) = 0.5 boundary, on its lower probability side. We note that T33 is predicted as a responder despite its position on the lower probability side of the displayed boundary, because its PD-L1 evasion activation contributed sufficiently to raise its response probability above 0.5. The remaining five pairwise concept spaces are presented in Supplementary Figure S3.

**Figure 4:**
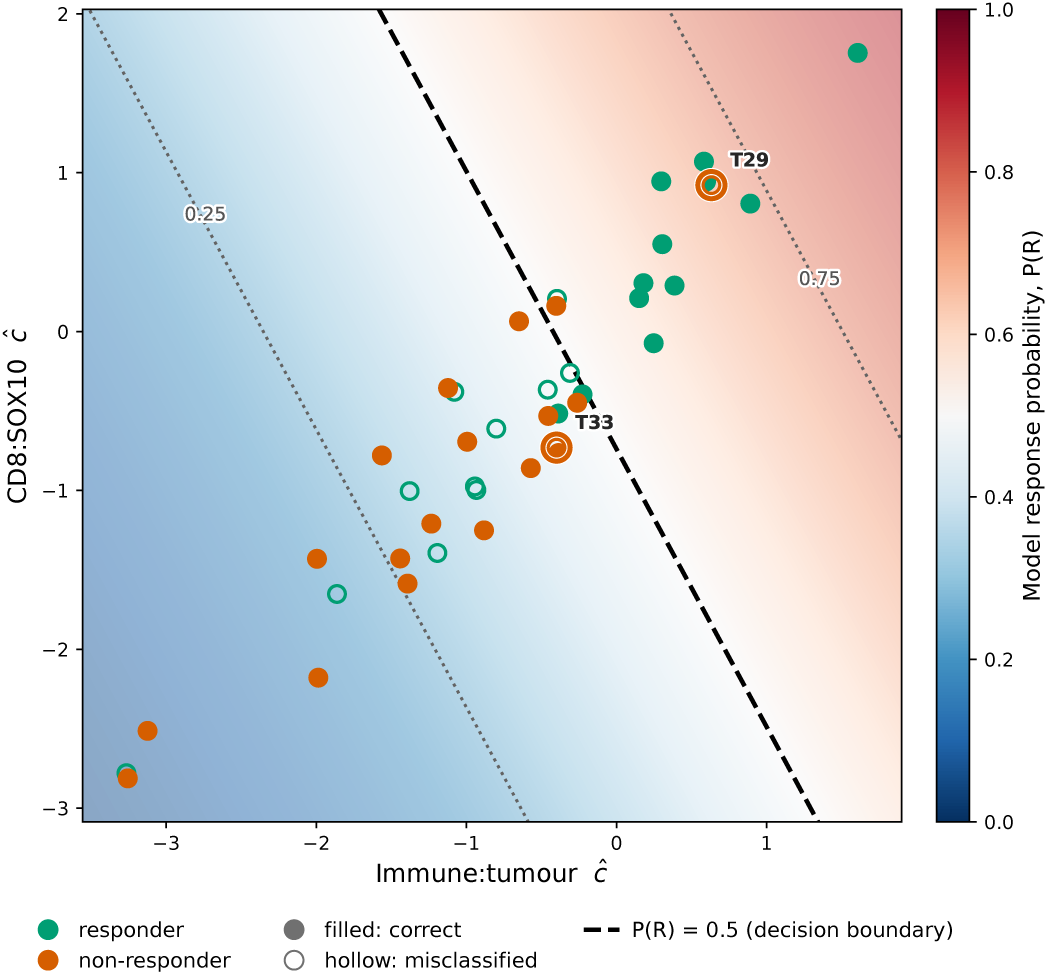
Joint concept decision space for the independent test cohort. Patients are positioned according to their target-scale learned activations for immune:tumour ratio and CD8:SOX10 balance. The background shows CB-COMIL’s predicted response probability, *P* (R), with CD68 relative signal and PD-L1 evasion fixed at their training cohort means. The dashed contour indicates *P* (R) = 0.5, while the dotted contours indicate *P* (R) = 0.25 and 0.75. Colours denote the true treatment outcome; filled markers indicate correct classifications and hollow markers indicate misclassifications based on all four patient-specific concept activations. T29 and T33, highlighted as orange hollow markers, are non-responders incorrectly predicted as responders. The displayed boundary is conditional on the fixed values of the other two concepts and therefore need not reproduce each patient’s full-model classification.

These analyses characterise how the learned concepts determine CB-COMIL’s output. Each response logit is exactly decomposed into four weighted concept contributions and a constant bias, with immune:tumour ratio and CD8/SOX10 balance generally contributing most strongly. This decomposition accounts for both correct and incorrect predictions, while the correspondence between learned activations and their intended biology is assessed separately.

### Measured biological concepts do not reproduce the predictive utility of learned concepts

Having characterised the concepts’ contributions to prediction, we next examined their correspondence with the intended biological quantities. We therefore assessed biological fidelity through two complementary analyses: comparing learned concept activations with independently measured mIF quantities and performing corrective interventions to determine whether measured values could replace learned activations in the prediction head.

Across the 41 patients in the independent test cohort with complete mIF data, all four learned concepts showed measurable correspondence with their independently quantified biological counterparts (Figure 5a–d). Spearman correlations were *ρ* = 0.61 for immune–tumour ratio, 0.66 for CD8/SOX10 balance, 0.76 for CD68 relative signal, and 0.72 for PD-L1 evasion. These positive correlations indicate that patients with higher measured values generally had higher learned activations, but do not establish numerical agreement, as Spearman correlation is unchanged by a constant shift or rescaling of the learned values. The deviations from the identity lines show that the learned activations do not reproduce the measured quantities exactly, so the concepts retain measurable biological alignment without functioning as direct measurements of their corresponding biomarkers.

**Figure 5:**
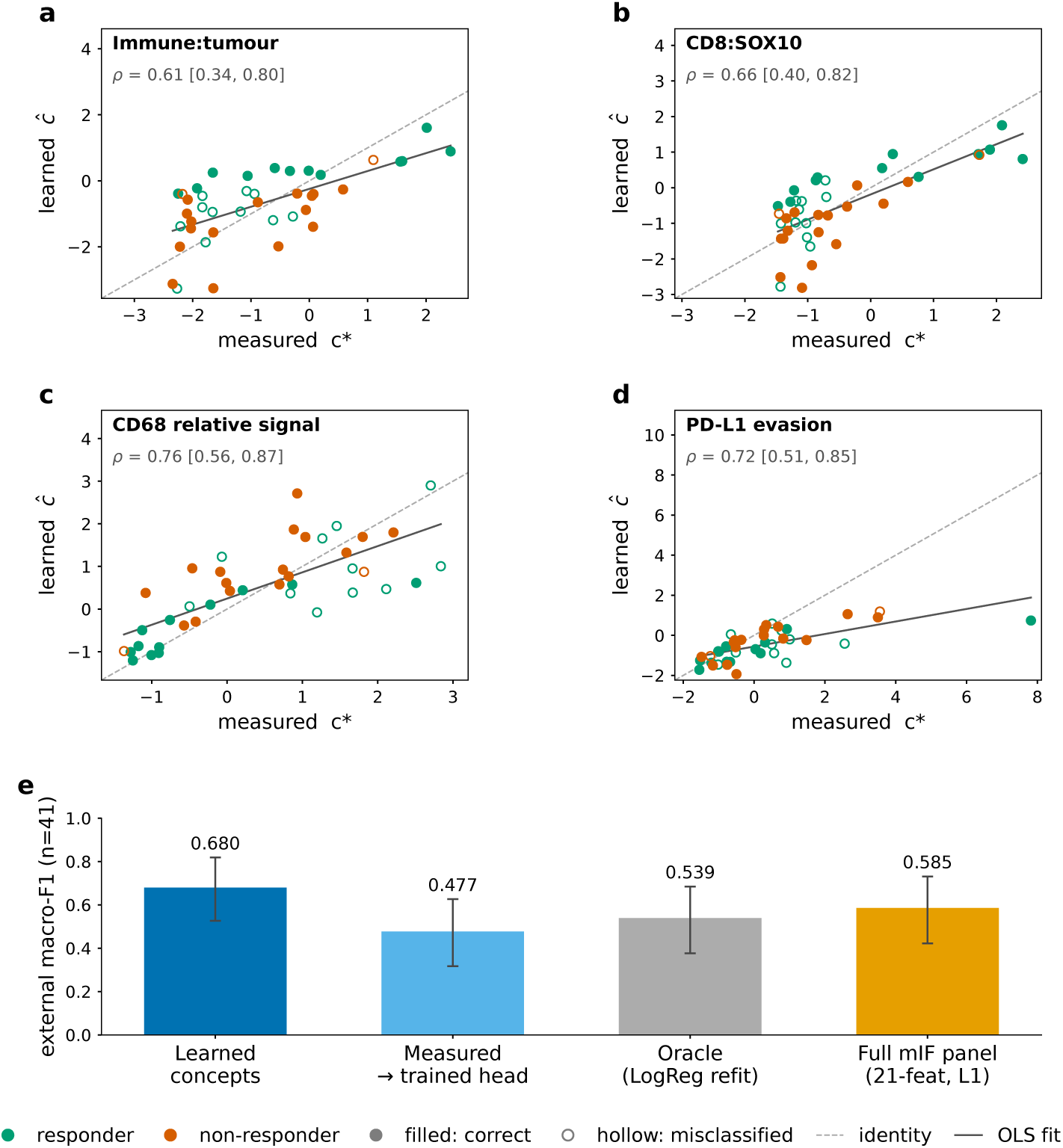
Fidelity and predictive utility of learned biological concepts in the independent test cohort. **a–d**, Measured and CB-COMIL learned concept values for the immune to tumour ratio, CD8:SOX10 balance, relative CD68 signal and PD-L1 evasion index, respectively, in 41 patients with complete mIF data in the independent test cohort. Measured values (*c^∗^*) were transformed using log(1 + *x*) and standardised using the training mean and standard deviation for each concept. CB-COMIL predicts learned values (*ĉ*) directly on this scale. Spearman’s *ρ* and its 95% confidence interval summarise the rank association for each concept. The dashed identity line indicates exact agreement, and the solid line shows the ordinary least squares fit. T29 and T33, two clinical non-responders classified as responders by the model, are highlighted in all four panels. **e**, Macro-F1 in the independent test cohort using the learned concepts; the measured concepts substituted into the trained prediction head; an oracle in which logistic regression was refitted on the four measured concepts; and an *L*_1_-regularised logistic regression on the full 21-feature measured panel. Bars show macro-F1, and error bars represent 95% percentile bootstrap confidence intervals obtained from 10,000 patient-level resamples with replacement of the 41 patients in the independent test cohort. Panels **a–d** assess biological correspondence, while panel **e** evaluates whether the measured concepts reproduce the predictive utility of the learned representation.

We tested whether the measured biological quantities could substitute for the learned concepts in predicting treatment response. Following the concept intervention framework of Koh et al. [21], we replaced all four learned activations with their independently measured mIF counterparts while keeping the trained prediction head unchanged. This substitution reduced macro-F1 from 0.68 to 0.48 among the 41 patients in the independent test cohort with complete mIF measurements (Figure 5e). Because the head was trained on learned activations, we also evaluated an oracle condition in which logistic regression was refitted using the measured concepts. Refitting yielded a macro-F1 of 0.54, partially recovering performance but remaining numerically below the learned-concept model.

To examine whether restricting the measured inputs to four summaries limited performance, we additionally evaluated an *L*_1_-regularised logistic regression using the full 21-feature handcrafted mIF panel (Table 2). This model achieved a macro-F1 of 0.59 on the independent test cohort, compared with 0.54 for the model using the four measured concepts and 0.68 for CB-COMIL (Supplementary Table S4). Expanding the measured feature set therefore yielded higher performance in this comparison, but did not match the learned-concept model. Both measured-feature models also performed worse on the test cohort than in cross-validation, whereas CB-COMIL did not (Supplementary Table S4), suggesting that staining-related shift in measured intensities may contribute to this gap.

**Table 2:** The 21-feature intensity-derived mIF panel used in the fusion ablation: six per-marker signal-to-noise ratios (SNR), nine derived immune ratios and indices, and six therapy- and markerinteraction terms. Marker symbols denote slide-level mean intensities; “therapy” is the treatment indicator (anti–PD-1 vs. anti–PD-1 + anti–CTLA-4).

| # | **Feature** | **Definition** |
|---|---|---|
| *Per-marker signal-to-noise ratios (SNR) (normalised by autofluorescence)* |  |  |
| 1 | CD8 SNR | CD8/AF |
| 2 | CD68 SNR | CD68/AF |
| 3 | CD16 SNR | CD16/AF |
| 4 | PD-L1 SNR | PD-L1/AF |
| 5 | SOX10 SNR | SOX10/AF |
| 6 | DAPI SNR | DAPI/AF |
| *Derived immune ratios and indices* |  |  |
| 7 | Immune–tumour ratio | (CD8 + CD16)/SOX10 |
| 8 | CD8/SOX10 balance | CD8/SOX10 |
| 9 | CD8/CD68 balance | CD8/CD68 |
| 10 | CD68 relative signal | CD68/(CD8 + CD68) |
| 11 | PD-L1 evasion index | PD-L1/CD8 |
| 12 | CD68:CD8 balance | (CD68 · PD-L1)/CD8 |
| 13 | Tumour-evasion proxy | SOX10 · PD-L1 |
| 14 | SOX10/DAPI ratio | SOX10/DAPI |
| 15 | CD16:CD68 balance | CD16 · (1 − CD68 relative signal) |
| *Therapy- and marker-interaction terms* |  |  |
| 16 | Therapy × PD-L1 evasion | therapy · (PD-L1/CD8) |
| 17 | Therapy × CD16 | therapy · CD16 |
| 18 | Therapy × myeloid suppression | therapy · [(CD68 · PD-L1)/CD8] |
| 19 | CD8 × PD-L1 | CD8 · PD-L1 |
| 20 | SOX10 × CD8 × therapy | SOX10 · CD8 · therapy |
| 21 | Therapy × immune–tumour | therapy · [(CD8 + CD16)/SOX10] |

These findings support the interpretation of CB-COMIL’s learned concepts as biologically aligned representations that explicitly contribute to its predictions. Their moderate to strong positive correlations with the corresponding measured biological targets indicate that higher measured values generally correspond to higher learned activations, but do not establish numerical equivalence. The reduction in predictive performance following concept substitution further indicates that the learned activations are not directly interchangeable with measured quantities and should not be interpreted as calibrated biomarker measurements.

### Ablation study

We evaluated the contributions of CB-COMIL’s principal components: marker-aware KRONOS features, a COMIL aggregator, and a concept bottleneck head, together with class-agnostic attention and instance-level supervision as further design choices (Supplementary Table S5). Across the evaluated configurations, cross-validation macro-F1 ranged from 0.62 to 0.70, whereas macro-F1 on the independent test cohort ranged from 0.38 to 0.69. The complete CB-COMIL configuration achieved the highest observed test macro-F1 (0.69).

Replacing marker-aware KRONOS with the global encoder reduced macro-F1 on the independent test cohort from 0.69 to 0.60, despite higher cross-validation macro-F1 (0.70 versus 0.67). Alternative aggregators also performed less well on the independent test cohort, particularly flat ABMIL (macroF1 of 0.38). Replacing the concept bottleneck with a linear prediction head applied directly to the 384-dimensional slide embedding removed both concept structure and concept supervision. On the independent test cohort, this reduced macro-F1 from 0.69 to 0.54, AUPRC from 0.77 to 0.64, and specificity from 0.90 to 0.47.

On the independent test cohort, CB-COMIL achieved a macro-F1 of 0.69, compared with 0.67 for opaque COMIL. The paired difference was +0.020 (95% bootstrap CI, *−*0.103 to +0.144, *p* = 0.76), with no statistically detectable difference in macro-F1 between the models. To assess robustness to random initialisation, we repeated training with three additional seeds. CB-COMIL and opaque COMIL showed competitive observed performance across the four runs, with macro-F1 ranging from 0.63 to 0.69 and 0.56 to 0.67, respectively. The paired differences ranged from +0.020 to +0.108, with all 95% bootstrap confidence intervals including zero (Supplementary Table S9). Removing instance-level supervision or replacing class-agnostic attention with per-class attention shifted the operating point, while AUPRC on the independent test cohort remained 0.77 at the reported precision. Without instance-level supervision, specificity fell to 0.42 and sensitivity rose to 0.78. With per-class attention, specificity was 1.00 and sensitivity was 0.13.

## Discussion

The value of AI predictions in cancer care extends beyond accuracy to understanding why a model favours treatment response or non-response for an individual patient. CB-COMIL addresses this need by expressing melanoma immunotherapy response predictions from mIF images through four learned representations of the tumour immune microenvironment: immune:tumour ratio, CD8/SOX10 balance, CD68 relative signal, and PD-L1 evasion. These representations, termed concepts, are trained to align with corresponding mIF-derived biological measurements. On the independent test cohort, CB-COMIL achieved comparable observed predictive performance to opaque COMIL across four random initialisations, while making individual predictions interpretable through the contributions of learned concepts. All four learned concepts showed moderate to strong positive correlations with their corresponding mIF-derived measurements in the independent test cohort (Figure 5), supporting their use as biologically aligned representations for examining the model’s predictions.

While the learned concepts provide a biologically informed basis for examining predictions, attention-based explanations offer a different perspective by identifying influential image regions. Removing the patches that attention ranked highest lowered macro-F1 more quickly than removing random patches (Fig. 2a), indicating that these patches influence the prediction. Retaining only the top-ranked patches, however, recovered performance no faster than retaining a random subset (Fig. 2b): even a small random fraction of the slide restored near-full performance, suggesting that multiple subsets of tissue can contain enough information to recover performance at the cohort level. Attention maps can therefore guide inspection of tissue regions that may inform the prediction, but do not establish which biological characteristics determine the prediction.

In contrast, CB-COMIL makes the contribution of each learned concept to the final prediction explicit. By constraining predictions to four learned concepts and a linear decision head, each response logit can be decomposed exactly into four signed concept contributions and a constant bias, without an alternative prediction pathway. Because the decision pathway is restricted to these concepts, the explanations reflect the model’s actual decision mechanism rather than a retrospective approximation. Concept interventions quantified the relative influence of each concept, with immune:tumour ratio and CD8/SOX10 balance exerting the greatest influence over the evaluated range (Table 1).

Patient-level decompositions could support clinical interpretation by revealing the biologically aligned learned representations underlying individual predictions, rather than providing a probability alone (Figure 3). In T19, a responder incorrectly predicted as a non-responder, low learned immune:tumour and CD8/SOX10 activations drove the prediction towards non-response, with no substantial opposing contribution from the other concepts. The same pattern was observed among the eleven missed responders, who had lower learned immune:tumour activations than correctly classified responders (median *−*0.94 versus +0.30), contributing to their misclassification as non-responders (Supplementary Table S2). Conversely, the model incorrectly classified T29, a clinical non-responder, as a responder. T29’s learned scores for the immune to tumour ratio and CD8/SOX10 balance were as high as those of correctly classified responders, helping to explain the model’s prediction. Understanding why T29 did not respond to treatment would require separate biological evidence. The decomposition illustrates the transparency of the decision process and the distinction between explaining a model’s prediction and explaining an individual patient’s treatment outcome. Further investigation could examine factors beyond the four concepts, including immune cell organisation and functional state. Decompositions of this kind could allow clinicians to see which tissue features drove a prediction and to judge when it warrants caution.

The usefulness of these explanations also depends on the choice of concepts. Leave-one-concept-out ablations showed that removing any single concept produced only modest changes in predictive performance, suggesting that the model does not rely critically on any one concept (Supplementary Table S6). Notably, removing CD68 relative signal produced the largest observed decrease in performance, despite its low weight in the prediction head (Table 1). This suggests that concept supervision may influence the shared representation beyond a concept’s direct contribution to the prediction. We nevertheless retained all four, because they were selected *a priori* for their biological relevance to checkpoint blockade, their measurable counterparts in the mIF panel, and their ability to represent immune–tumour relationships compactly. Together, they provide a compact, biologically motivated basis for interpreting model predictions, although their individual contributions may overlap.

Importantly, biological correspondence did not necessarily translate into predictive importance. CD68 relative signal had the highest observed correlation with its measured counterpart (*ρ* = 0.76) (Figure 5), yet showed the smallest intervention effect (Table 1) and generally the smallest contributions to individual predictions (Fig. 3). Its limited influence could reflect information shared with other concepts or limitations of the selected summary, rather than a lack of biological relevance. Likewise, the positive prediction weight for learned PD-L1 evasion describes its contribution when the other concepts are held fixed (Table 1); it does not establish that immune evasion promotes treatment response. These contrasts show that a concept’s contribution to the model’s prediction does not directly establish its biological role in treatment response.

Correlation with a measured quantity does not make a learned concept a substitute for it. Replacing the learned activations with their measured counterparts in the trained prediction head reduced macro-F1 from 0.68 to 0.48, while refitting the prediction head on measured concepts achieved 0.54 (Figure 5e). A model trained on the full panel of 21 measured mIF features achieved a macro-F1 of 0.59 on the independent test cohort. Neither intermediate nor late fusion of measured features with image embeddings exceeded the macro-F1 of opaque COMIL (Supplementary Table S4). These findings suggest that learned concepts may retain predictive information not captured by the selected measured summaries, while adding those measurements to image embeddings provided no improvement in the evaluated fusion configurations. Two mechanisms may contribute: concept leakage under joint training, in which the response objective encodes information beyond the measured targets in the learned concepts [25, 26], and staining-related shift in measured intensities between cohorts. Thus, although the learned concepts support interpretation of the model’s predictions, their numerical values should not be treated as calibrated biological measurements or substitutes for laboratory assay results.

For precision oncology, identifying patients unlikely to respond to immunotherapy could help prioritise further clinical assessment and consideration of alternative treatment strategies. At the evaluated threshold, CB-COMIL correctly identified more non-responders than opaque COMIL (17/19 versus 13/19), but fewer responders (12/23 versus 15/23). CB-COMIL predicted non-response for 28 patients, 11 of whom responded to immunotherapy. The proportion of non-response predictions that were correct was therefore similar for CB-COMIL and opaque COMIL (61% versus 62%, respectively), despite a numerically higher AUPRC of CB-COMIL (0.773 versus 0.702). The ablation results showed a similar distinction between performance across thresholds and performance at a selected threshold, with comparable AUPRC values yielding markedly different recall for responders and non-responders. At the evaluated threshold, the number of responders misclassified as non-responders argues against using the model to withhold immunotherapy. Clinical use would therefore require assessment of calibration and selection of a decision threshold that accounts for the clinical consequences of prediction errors.

The independent test cohort comprised 42 patients, of whom 41 had complete biological measurements. This cohort provides a valuable independent evaluation, as mIF datasets are costly and technically demanding to generate, and comparable public cohorts with treatment response annotations are scarce. Notably, the independent test cohort was acquired using different imaging instrumentation and staining protocols and exhibited statistically significant differences in marker signal distributions relative to the development cohort, including within responder and non-responder groups (two-sided Mann–Whitney U test; Supplementary Fig. S7). These differences provide quantitative evidence of domain shift, potentially reflecting variation in both acquisition conditions and case mix between cohorts, and allow CB-COMIL to be evaluated under conditions different from those used for model development. Patient-level bootstrap confidence intervals and evaluations across four random seeds were used to assess sampling uncertainty and sensitivity to training variability, respectively (Supplementary Table S9). Nevertheless, validation in larger independent cohorts remains necessary to establish broader generalisability.

The concept targets are derived from slide-level mean marker intensities, capturing relative marker signals rather than the spatial organisation of cells within the tumour microenvironment. These intensity-based ratios provide biologically motivated supervision at the same level as the aggregated slide representation used to predict the concepts. Spatial features, including the proximity of CD8 T cells to tumour cells, have also been associated with checkpoint blockade response [6]. Future work could extend the concept set to include cell densities, immune cell phenotypes and spatially defined tumour–immune interactions.

CB-COMIL makes melanoma immunotherapy response predictions from mIF whole-slide images inspectable through four predefined, biologically supervised concepts. On the independent test cohort, it made each concept’s contribution explicit while achieving comparable observed macro-F1 to the opaque comparator. The learned concepts correlated with their measured biological targets, but substituting those targets into the trained prediction head reduced performance. Together, these findings show the value of assessing both how each concept contributes to a prediction and how closely its learned values correspond to tissue measurements.

## Methods

### Dataset

The model is trained using 211 slide-level labelled whole-slide mIF images of melanoma tissue sourced from the MIA. A separate set of 42 images was processed using a different staining sequence and imaging instrument, introducing potential technical distribution shifts in fluorescence signals and providing an opportunity to assess model generalisability across different acquisition conditions. No patient contributed slides to both the development and test cohorts. Individual images reach up to 24,000 *×* 24,000 pixels at a resolution of 0.5 *µ*m per pixel. Each image comprises five protein biomarker channels (CD8, CD16, CD68, PD-L1 and SOX10), together with a nuclear stain (DAPI) and an autofluorescence (AF) channel. All patients received anti-PD-1 monotherapy or combined anti-PD-1 and anti-CTLA-4 therapy. Tissue samples were labelled according to 6-month progression-free survival, with non-responsive cases defined as the negative class (91 training and 19 test samples) and responsive cases as the positive class (120 training and 23 test samples). Biopsies were collected mainly within one year before treatment (89%), with a limited number taken more than a year before therapy. Full acquisition details appear in [7, 27].

### Participants and specimens

Patients with unresectable stage III or stage IV melanoma who had received or were eligible for anti-PD-1 monotherapy (PD-1) or combined anti-PD-1 and anti-CTLA-4 immunotherapy (IPI+PD-1) were eligible for inclusion. Melanoma subtypes included cutaneous, mucosal, acral, subungual and melanoma of unknown primary. Patients with uveal melanoma, although included in prospective biomarker testing within the broader PIP-PREDICT programme, were excluded from the present analysis because of its distinct biological and clinical characteristics. Prior adjuvant or neoadjuvant systemic therapy was permitted.

Pretreatment formalin-fixed, paraffin-embedded (FFPE) tumour specimens were obtained from the Melanoma Institute Australia Biobank. Specimens containing fewer than 100 melanoma cells were excluded. The study was conducted in accordance with the Declaration of Helsinki and approved by the Sydney Local Health District Human Research Ethics Committee (2020/ETH00426).

### Response classification

Treatment response was defined using progression-free survival (PFS) measured from immunotherapy initiation. Patients who experienced disease progression within six months of treatment initiation (PFS *<* 6 months) were classified as non-responders, whereas those with PFS *≥* 6 months were classified as responders.

### Concept-target data

The four concept targets used to supervise the bottleneck are not annotations but quantities measured directly from the same mIF panel that produced the images, so each is independently re-measurable in a pathology laboratory. For slides with complete HALO measurements, the seven channels were quantified to slide-level mean intensities in HALO (Indica Labs) image-analysis software, yielding one mean intensity per channel: the five biomarkers on their respective Opal fluorophores (CD8 Opal570 Intensity, CD68 Opal520 Intensity, CD16 Opal620 Intensity, PDL1 Opal650 Intensity, SOX10 Opal690 Intensity), the nuclear channel (DAPI Intensity), and autofluorescence (Autofluorescence Intensity). From these HALO intensities we first formed per-marker signal-to-noise ratios by normalising each marker to autofluorescence (marker SNR = marker*/*AF). The four concepts were then computed as composite ratios of these SNRs following their biological definitions (Table 2): the immune–tumour ratio (CD8 + CD16)*/*SOX10, the CD8/SOX10 balance, CD68 relative signal CD68*/*(CD8 +CD68), and the PD-L1 evasion index PD-L1*/*CD8. The slide-level concept values serve as supervision targets during training, rather than as model inputs. CB-COMIL learns to predict these concepts from KRONOS image features, allowing response predictions without requiring measured concept values at inference. Concept targets were available for 181 training and 41 test slides with a complete quantified mIF panel. The remaining 30 training slides lacked concept targets and contributed to the response and instance-level losses, but not to the concept loss. The single test slide with missing intensities was evaluated using image-derived features alone. Because the ratios are right-skewed, each concept was log(1 + *x*)-stabilised and then *z*-scored; to prevent leakage, the standardisation statistics were estimated on the complete-mIF training slides of each fold only and applied unchanged to validation and test, with the final retrained model using statistics from all complete training slides.

### Concept Bottleneck COMIL (CB-COMIL)

Concept Bottleneck COMIL (CB-COMIL) is built on our previously developed Channel Optimisation with Multi Instance Learning (COMIL) framework for whole-slide multiplex immunofluorescence [9], which aggregates per-marker patch features through a gated channel transform and gated-attention pooling to a slide-level prediction. We extend COMIL by incorporating marker-aware KRONOS features and replacing its conventional classifier with a concept bottleneck that routes predictions through four predefined immune concepts. These modifications retain marker-specific information and make the contribution of each learned concept to the final prediction explicit. The following sections describe the preprocessing, feature extraction, aggregation, concept bottleneck, training objective and reference models.

#### Preprocessing

Whole-slide seven-channel mIF images were stored as multi-resolution pyramidal TIFFs. Every channel was resolved to its marker by name from per-slide metadata before any pixel was read.

A tissue foreground mask was computed from the union of the DAPI, CD8, and PD-L1 channels [9]. Following COMIL, each slide was then partitioned at a resolution of 0.5 *µ*m/px (*≈* 20*×*) into non-overlapping 224 *×* 224 px patches, corresponding to an approximately 112 *µ*m field of view. A patch was retained if at least 25% of its pixels fell within the tissue mask.

#### Feature extraction and aggregation

The retained 224-px patches were encoded with the pretrained KRONOS[28] pathology foundation model after a two-stage normalisation: per-slide, per-channel intensity rescaling to [0, 1] using the 0.5th/99.5th percentiles (computed once per slide over a pixel sample), followed by KRONOS’s fixed per-marker mean/std standardisation. We adopted a marker-aware configuration, in which each of the six retained channels (DAPI, SOX10, CD8, CD68, CD16, and PD-L1), with autofluorescence (AF) excluded, was encoded independently as a single-channel input, yielding six 384-dimensional per-marker feature representations per patch. KRONOS identifies each input channel by a marker token: the five biomarkers and DAPI were mapped to their native KRONOS tokens, while SOX10 was mapped to the S100 melanocyte-lineage token as the closest available proxy. AF, having no corresponding token and representing background signal rather than a stained marker, was excluded, leaving the six channels above.

Patch-level features were aggregated to a slide-level prediction using COMIL [9]. Each whole-slide mIF image with *C* channels is represented as a bag of *N_p_* patches, where each patch is a *C ×* 384 tensor of per-marker KRONOS features. Following COMIL, aggregation proceeds in three stages: a gated channel transformation (GCT) [29] that recalibrates the marker channels according to their multiplex context, attention-based pooling that weights patches by their relevance to the slide-level label and collapses the bag to a slide embedding, and an instance-level clustering objective that learns class-discriminative patches during training.

##### Gated channel transformation

The GCT comprises three components: global context embedding, channel normalisation, and gating adaptation. CB-COMIL operates on 384-dimensional (D=384) per-marker KRONOS features. For patch *n*, let **f***_n,c_ ∈* R^384^ denote the feature vector of channel *c ∈ {*1*, . . . , C}*.

*Global context embedding.* Each channel’s global context is embedded as its *L*_2_ feature magnitude under a learnable embedding weight *α_c_*,

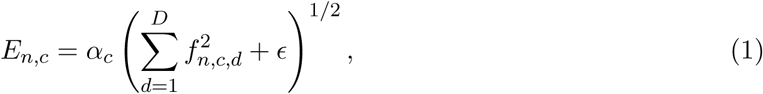

where *ɛ* is a small constant for numerical stability and *α_c_* adjusts the significance of each channel; a low *α_c_* removes channel *c* from the subsequent normalisation, allowing the module to adapt to each marker’s characteristics.

*Channel normalisation.* The channel embeddings are normalised across the *C* channels by *L*_2_ normalisation,

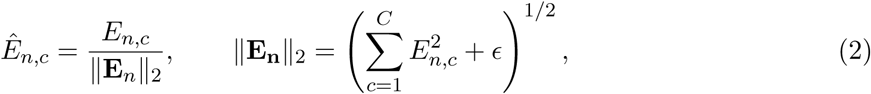

so that each channel embedding reflects its relative contribution among all the markers. *Gating adaptation.* A gate is formed from the normalised context through a learnable per-channel gating weight *γ_c_* and bias *β_c_*, and the channel features are rescaled by it,

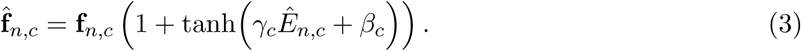

A positive *γ_c_* amplifies a channel’s activations, promoting competition and emphasising the most discriminative markers, whereas a negative *γ_c_* suppresses strong activations and relatively boosts weaker ones, fostering cooperation; the 1 + tanh(*·*) form models identity mapping and prevents vanishing gradients during training. This lets the model up- or down-weight individual markers according to each patch’s overall multiplex context rather than treating channels independently.

Following gating adaptation, the gated channels are collapsed by average pooling across the six channels, yielding a single patch embedding,

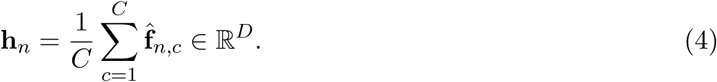

##### Attention mechanism

Because labels are available only at the slide level, the set of patch embeddings 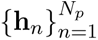 is aggregated into a single slide representation using attention-based pooling [30]. The attention mechanism learns which patches are most informative for predicting the slide-level label by assigning each patch a scalar attention weight. Specifically, attention is computed using a gated two-branch network, in which a tanh branch encodes the patch representation and a sigmoid branch gates it,

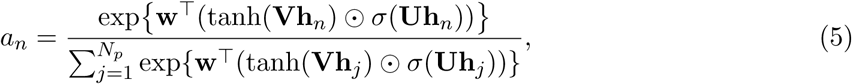

where **U**, **V** *∈* R*^L×D^* and **w** *∈* R*^L^* are learnable parameters, L=256 is the attention hidden dimension, *σ*(*·*) denotes the sigmoid activation, *⊙* denotes the element-wise product, and the attention weights are normalised across patches using a softmax function. Unlike the original COMIL, which learned separate attention distributions for each class, CB-COMIL uses a single, class-agnostic attention distribution for binary response prediction. The resulting slide embedding is computed as the attention-weighted sum of the patch embeddings,

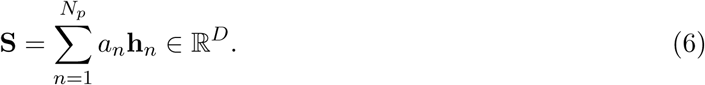

##### Instance-level clustering

To learn class-discriminative patches, COMIL’s instance-clustering objective was retained. At each training step, the *B* highest- and lowest-attention patches were pseudo-labelled as supporting and opposing evidence, respectively, and classified using an auxiliary linear instance-level head trained with cross-entropy loss. This instance-level objective complements the slide-level objective by supervising local features. The two objectives are combined in the overall training loss using a fixed weight, *µ*, for the instance clustering objective (see the Training objective).

#### Concept bottleneck head

To enable interpretability by construction, the response prediction is routed through a small set of named, biologically meaningful concepts. These were pre-specified from the established immunobiology of immune-checkpoint blockade (ICB) response and from the quantities measurable on the mIF panel, before any association with clinical outcome was examined. This design choice follows the principle that concept bottlenecks are defined by human-specified concepts [21]; selecting concepts based on outcome associations would reintroduce the opacity that the bottleneck is designed to remove and risk overfitting to the development cohort.

The four CB-COMIL concepts were selected according to four criteria. First, each concept is an intensity-based proxy computed from slide-level mean marker intensities and maps to a canonical biological axis of immune checkpoint blockade (ICB) response. The *immune–tumour ratio* approximates the abundance of immune signal relative to tumour signal, reflecting the degree of immune infiltration. The CD8/SOX10 balance approximates cytotoxic T-cell signal relative to melanoma-lineage signal, indicating whether the effector population that checkpoint blockade reactivates is present in proportion to the tumour. CD68 relative signal approximates the share of macrophage signal within the CD8 and CD68 compartment; macrophages can suppress cytotoxic activity, although their association with response depends on phenotype [8]. The PD-L1 evasion index approximates PD-L1 signal relative to CD8 signal, a proxy for the balance between checkpoint-associated and cytotoxic signals. Second, each concept has a measurable biological counterpart that can be independently quantified from the same mIF panel. This provides an external biological reference for evaluating the learned concept representation. Third, each concept is defined as a composite ratio rather than a single raw marker measurement, enabling representation of interpretable biological relationships rather than isolated marker abundance. Fourth, the concept set is intentionally restricted to four quantities, so that the bottleneck remains a genuine biological abstraction rather than becoming another high-dimensional latent representation. Consequently, every prediction must be expressed through these four predefined biological axes.

These four concepts are realised architecturally as a bottleneck layer. Following feature aggregation, the conventional classifier head is replaced with a two-layer prediction head comprising a concept layer and a linear response layer. The *D*-dimensional slide embedding **S** is first projected onto *K* = 4 interpretable concepts, which are subsequently mapped to the response logit through a linear layer,

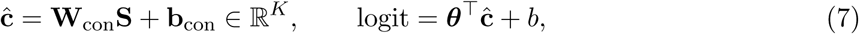

where **W**_con_ *∈* R*^K×D^*, **b**_con_ *∈* R*^K^*, ***θ*** *∈* R*^K^* and *b ∈* R are learnable parameters. Because the response layer is linear, the prediction logit decomposes exactly into a baseline bias *b* and *K* additive signed contributions, *θ_k_ĉ_k_*, each corresponding to a learned concept.

##### Concept targets

The measured concept values used to supervise **ĉ** are right-skewed ratios, and were therefore log(1+*x*)-stabilised and then *z*-scored so that the four concepts contribute comparably to the concept loss. To prevent information leakage from held-out slides, the standardisation statistics were estimated on the complete-mIF training slides of each fold only and applied unchanged to validation and test; the final retrained model used statistics from all complete training slides.

#### Training objective

CB-COMIL is trained end-to-end with a three-term loss,

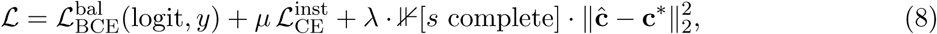

where 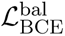 is a class-balanced binary cross-entropy (BCE) on the response, 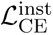 is COMIL’s instance-clustering loss weighted by a fixed *µ*, and the third term supervises the bottleneck towards the standardised true concepts **c***^∗^* with weight *λ*. This concept term is gated by a completeness indicator ⊮[*s* complete]: only slides with a complete mIF panel contribute to it, whereas every slide, including the 30 training and 1 test cases without quantified mIF, contributes fully to the response and instance terms. Missing-modality slides therefore still train the backbone and response head but exert no pull on the concept layer, so the bottleneck is never supervised by imputed or fabricated biology. The instance weight *µ* weights instance-clustering loss, whereas the concept weight *λ* controls the strength of concept supervision. Both weights were selected by five-fold cross-validation on the development cohort (see Training).

##### Training

Models were trained per bag (batch size 1) for 50 epochs. For CB-COMIL, the objective combined the balanced BCE with the concept-supervision term 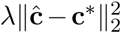, applied only to complete-mIF slides, and the instance-clustering loss with *B* = 8. The instance-loss weight *µ* was selected over *{*0, 0.1, 0.2, 0.3, 0.5, 0.7*}*, yielding *µ* = 0.3.

The concept-supervision weight *λ* was evaluated on the development cohort by five-fold crossvalidation over *{*0, 0.3, 0.5, 0.7, 0.8*}* (Supplementary Fig. S8). Cross-validation macro-F1 varied little across non-zero values (0.668–0.686), and all values lay within one fold standard deviation of the unsupervised setting (*λ* = 0: 0.682 *±* 0.061). Concept fidelity, measured as the mean per-concept *R*^2^ between learned and standardised measured concepts on complete-mIF validation slides, increased with *λ*, from 0.40 *±* 0.18 at *λ* = 0.3 to 0.52 *±* 0.07 at *λ* = 0.8. *R*^2^ was used here because it penalises scale and offset error, whereas the rank correlation reported for the independent test cohort assesses ordering only. We selected the smallest non-zero weight (*λ* = 0.3) to introduce explicit concept supervision while limiting its contribution to the training objective.

Following COMIL, the embedding weight *α* was initialised to 1 and the gating weights and biases to 0. Optimisation used Adam (learning rate 10*^−^*^3^, weight decay 10*^−^*^4^). Model selection used stratified five-fold cross-validation. The epoch selected within each fold was recorded, and the five selected epochs were averaged and rounded to obtain a single retraining epoch. The final model was then retrained on all 211 development whole-slide mIF images for this number of epochs and evaluated once on the 42-slide independent test cohort. All results reported in the main text use a fixed seed of 42. To assess robustness to initialisation, we repeated training with three further seeds (0, 1, 2) under identical configuration, with no per-seed tuning and the retraining epoch recomputed by the same procedure, and report the resulting macro-F1 and concept fidelity in Supplementary Table S9.

Point estimates on the independent test cohort are reported with 95% percentile bootstrap confidence intervals (*B* = 10,000, patients resampled with replacement). Pairwise model comparisons use a paired bootstrap on the macro-F1 difference, with significance declared when the 95% confidence interval of the difference excludes zero. The *ORACLE* (*L*_2_-regularised logistic model fit on the standardised true concepts) and full-panel (*L*_1_-regularised logistic model fit on the 21-feature mIF panel) comparisons are restricted to the 41 complete-mIF test patients.

### Relationship to the original COMIL

CB-COMIL preserves COMIL’s core design—the global-context channel gate, the attention mechanism, and instance-level clustering—but differs in four respects dictated by the encoder and task. First, the two models construct their per-marker inputs differently. Because ResNet101 requires a three-channel input, COMIL formed a pseudo-RGB image for each biomarker by stacking it with the DAPI and AF channels to supply morphological context; CB-COMIL instead exploits the marker-aware design of KRONOS, which encodes a single-channel input identified by a marker token, so each marker is encoded individually without auxiliary stacking. Second, because KRONOS features are 384-dimensional, we omit COMIL’s per-instance projection (2048 *→* 1024) and perform the gating, pooling, and aggregation in the native 384-dimensional space. Third, CB-COMIL uses a single, class-agnostic attention distribution rather than the per-class attention of the original COMIL. The present model operates on 384-dimensional KRONOS features coupled to a concept bottleneck head, where a single patch-importance map aligns directly with the concept-level interpretation reducing the attention parameter count. Fourth, the slide-level (BCE) and instance-clustering (cross-entropy) losses are combined using a fixed weight of 0.3 for the instance term, rather than the adaptive loss (ADL) used in the original COMIL [9]. This modification is particularly important for CB–COMIL, which introduces an additional concept supervision objective controlled by *λ*. Fixing the instance loss contribution avoids simultaneous dynamic reweighting of multiple objectives, allowing *λ* to provide an explicit and interpretable control over the tradeoff between predictive performance and concept fidelity.

### Attention attribution and faithfulness evaluation

We evaluated the faithfulness of patch-level attention using complementary deletion and insertion experiments on the independent test cohort (*n* = 42). Three patch-ranking strategies were compared: the attention weights produced by CB-COMIL, Integrated Gradients (IG), and a random ordering. IG provided an independent gradient-based attribution reference, while random ranking controlled for changes in predictive performance caused by the perturbation procedure itself.

#### Integrated Gradients

Attribution methods based solely on the local gradient can be affected by gradient saturation, whereby the gradient approaches zero even for features that contributed substantially to the model’s prediction. IG addresses this limitation by integrating gradients along a straight-line path from a reference input *x^′^* (the zero vector in our implementation) to the observed input *x*. For a differentiable model *F* : *X →* R, the attribution to feature *i* is

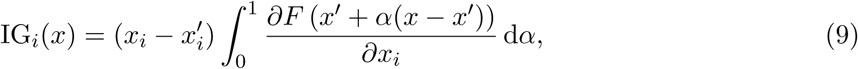

where (*x_i_ − x^′^_i_*) represents the difference between the observed and reference feature values, and the integral captures the model’s average sensitivity to that feature along the integration path. IG satisfies the completeness property,

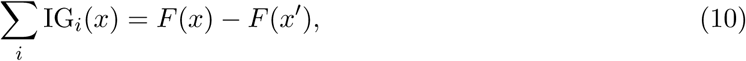

such that the feature attributions collectively account for the change in model output between the reference and observed inputs, up to numerical integration error. IG was computed with respect to the response logit, using 50 integration steps. Attributions were computed on the patch-level KRONOS features and summed over all feature dimensions and marker channels of each patch to give a single patch score, and patches were ranked by absolute score.

### Deletion and insertion experiments

We assessed two complementary properties of the attribution rankings, necessity and sufficiency, using deletion and insertion metrics [23]. Necessity was evaluated through deletion, in which patches were progressively removed in descending order of their attribution scores. Predictive performance was recalculated after each perturbation. A faster decline in performance indicates greater dependence on the highest-ranked patches.

Sufficiency was evaluated through insertion, in which patches were progressively retained in descending order of attribution, and predictive performance was assessed as additional patches were introduced. Patches were removed from the bag rather than masked in the image, and the attention weights were renormalised over the remaining patches. Rankings were computed once on the full bag and not updated after each step. Fractions from 0 to 0.9 were evaluated in steps of 0.05 for deletion, and from 0 to 1.0 for insertion, and areas under the curves were normalised by the evaluated fraction range. The random control was repeated over 10 random permutations, and its curves and areas are reported as the mean across permutations. Faster recovery of predictive performance provides stronger evidence that the highest-ranked patches retain information required for prediction.

Both experiments were performed using attention, IG, and random patch rankings. The random control underwent the same perturbation procedure as the attribution-based rankings.

### Faithfulness metrics

Deletion and insertion curves were summarised using the area under the curve (AUC) for macro-F1, non-responder specificity, non-responder F1, and area under the precision–recall curve (AUPRC). Lower deletion AUC indicates greater sensitivity to the removal of highly ranked patches, whereas higher insertion AUC indicates faster recovery of predictive performance.

We additionally evaluated prediction flip rate to quantify changes in predicted class following patch perturbation. Together, these metrics allowed us to assess whether highly ranked patches were influential in the model’s predictions and whether retaining them was sufficient to recover predictive performance.

## Data availability

The mIF images and associated clinical data are not publicly available owing to patient privacy restrictions. De-identified data may be made available from the corresponding author on reasonable request, subject to approval by Melanoma Institute Australia and the Sydney Local Health District Human Research Ethics Committee.

## Code availability

The source code for CB-COMIL, including model training and evaluation scripts, will be made publicly available upon publication.

## Ethics approval and consent to participate

Informed consent was obtained from all participants, and the study was conducted in accordance with approved ethical guidelines. Ethical approval was granted by the Sydney Local Health District Human Research Ethics Committee (protocol numbers: X17-0312 and 2019/ETH07604; reference numbers: 2020/ETH00426 and X20-0086). The clinical study, Personalised Immunotherapy Platform (PIP-PREDICT), is registered with ClinicalTrials.gov (NCT06536257).

## Author contributions

P.R. conceived the study, developed the methodology and software, performed the analysis and validation, prepared the figures and wrote the original draft. T.N.G. curated the data and provided resources. N.A.A., Y.M. and S.N.L. curated the data. S.B. contributed to interpretation of the results. E.C. acquired funding and contributed to interpretation of the results. G.V.L. provided resources and acquired funding. J.S.W. curated the data, provided resources, acquired funding and supervised the study. S.L. provided resources, including arranging data access, contributed to the study design and interpretation of the results, and supervised the study. All authors reviewed and edited the manuscript and approved the final version.

## Acknowledgements

This study was supported by the National Health and Medical Research Council (NHMRC) Centre for Research Excellence in Digital Health, an NHMRC Investigator Grant (GNT2008645), the Medical Research Future Fund (MRFF) National Critical Research Infrastructure project National Infrastructure for federated learNing in DigitAl Health (NINA; MRFCRI000246), and in part by the Australian Melanoma Research Foundation (AMRF). This work was also supported by Melanoma Institute Australia, the New South Wales Department of Health, NSW Health Pathology, the NHMRC and the Cancer Institute NSW (CINSW). Data generation was funded by an NHMRC Fellowship (J.S.W., APP1174325), a Melanoma Research Alliance Young Investigator Fellowship (700455), a Cancer Council NSW project grant (RG19-15) and a CINSW Translational Program Grant (TPG 2021/TPG2114). T.N.G. is supported by an NHMRC Investigator Grant (GNT2033462) and a CINSW Fellowship (2020/ECF1244). G.V.L. is supported by NHMRC Investigator Grants (GNT2007839 and GNT2018514) and by the Melanoma Foundation of the University of Sydney through the University of Sydney Medical Foundation. The funders had no role in study design, data analysis, interpretation or the decision to publish.

We thank the patients of Melanoma Institute Australia (MIA) and their families for their contributions to this study. We also thank colleagues at MIA, the Charles Perkins Centre (University of Sydney) and Royal Prince Alfred Hospital, and acknowledge support from the ClearBridge Foundation. This research was undertaken with the assistance of resources and services from the National Computational Infrastructure (NCI), which is supported by the Australian Government. We thank Satya Vedantam and Richard Miller for facilitating access.

## References

[1] Freddie Bray, Mathieu Laversanne, Hyuna Sung, Jacques Ferlay, Rebecca L Siegel, Isabelle Soerjomataram, and Ahmedin Jemal. Global cancer statistics 2022: Globocan estimates of incidence and mortality worldwide for 36 cancers in 185 countries. CA: A cancer journal for clinicians, 74(3):229–263, 2024.

[2] Xuwen Lin, Chenyu Zong, Zhihan Zhang, Weiyi Fang, and Ping Xu. Progresses in biomarkers for cancer immunotherapy. MedComm, 4(5):e387, 2023.

[3] Jedd D Wolchok, Vanna Chiarion-Sileni, Piotr Rutkowski, C Lance Cowey, Dirk Schadendorf, John Wagstaff, Paola Queirolo, Reinhard Dummer, Marcus O Butler, Andrew G Hill, et al. Final, 10-year outcomes with nivolumab plus ipilimumab in advanced melanoma. The New England journal of medicine, 392(1):11–22, 2025.

[4] Steve Lu, Julie E Stein, David L Rimm, Daphne W Wang, J Michael Bell, Douglas B Johnson, Jeffrey A Sosman, Kurt A Schalper, Robert A Anders, Hao Wang, et al. Comparison of biomarker modalities for predicting response to pd-1/pd-l1 checkpoint blockade: a systematic review and meta-analysis. JAMA oncology, 5(8):1195–1204, 2019.

[5] Pok Fai Wong, Wei Wei, James W Smithy, Balazs Acs, Maria I Toki, Kim RM Blenman, Daniel Zelterman, Harriet M Kluger, and David L Rimm. Multiplex quantitative analysis of tumor-infiltrating lymphocytes and immunotherapy outcome in metastatic melanoma. Clinical Cancer Research, 25(8):2442–2449, 2019.

[6] Tuba N Gide, Ines P Silva, Camelia Quek, Tasnia Ahmed, Alexander M Menzies, Matteo S Carlino, Robyn PM Saw, John F Thompson, Marcel Batten, Georgina V Long, et al. Close proximity of immune and tumor cells underlies response to anti-pd-1 based therapies in metastatic melanoma patients. Oncoimmunology, 9(1):1659093, 2020.

[7] Zarwa Yaseen, Tuba N Gide, Jordan W Conway, Alison J Potter, Camelia Quek, Angela M Hong, Georgina V Long, Richard A Scolyer, and James S Wilmott. Validation of an accurate automated multiplex immunofluorescence method for immuno-profiling melanoma. Frontiers in molecular biosciences, 9:810858, 2022.

[8] Hansol Lee, Angela L Ferguson, Camelia Quek, Ismael A Vergara, Ines Pires daSilva, Ruth Allen, Tuba Nur Gide, Jordan W Conway, Lambros T Koufariotis, Nicholas K Hayward, et al. Intratumoral cd16+ macrophages are associated with clinical outcomes of patients with metastatic melanoma treated with combination anti-pd-1 and anti-ctla-4 therapy. Clinical Cancer Research, 29(13):2513–2524, 2023.

[9] Priyanka Rana, Tuba N. Gide, Nurudeen A. Adegoke, Yizhe Mao, Shlomo Berkovsky, Enrico Coiera, James S. Wilmott, and Sidong Liu. Deep learning framework for classifying whole-slide multiplex immunofluorescence images to predict immunotherapy response in melanoma patients. IEEE Journal of Biomedical and Health Informatics, 30(10):8342–8353, 2026.

[10] Daniel Jiménez-Sánchez, Mikel Ariz, Hang Chang, Xavier Matias-Guiu, Carlos E de Andrea, and Carlos Ortiz-de Solorzano. Naronet: Discovery of tumor microenvironment elements from highly multiplexed images. Medical image analysis, 78:102384, 2022.

[11] Daniel Jiménez-Sánchez, Álvaro López-Janeiro, María Villalba-Esparza, Mikel Ariz, Ece Kadioglu, Ivan Masetto, Virginie Goubert, Maria D Lozano, Ignacio Melero, David Hardisson, et al. Weakly supervised deep learning to predict recurrence in low-grade endometrial cancer from multiplexed immunofluorescence images. NPJ digital medicine, 6(1):48, 2023.

[12] Weina Jin, Xiaoxiao Li, Mostafa Fatehi, and Ghassan Hamarneh. Guidelines and evaluation of clinical explainable ai in medical image analysis. Medical image analysis, 84:102684, 2023.

[13] Grégoire Montavon, Alexander Binder, Sebastian Lapuschkin, Wojciech Samek, and Klaus-Robert Müller. Layer-wise relevance propagation: an overview. Explainable AI: interpreting, explaining and visualizing deep learning, pages 193–209, 2019.

[14] Bolei Zhou, Aditya Khosla, Agata Lapedriza, Aude Oliva, and Antonio Torralba. Learning deep features for discriminative localization. In Proceedings of the IEEE conference on computer vision and pattern recognition, pages 2921–2929, 2016.

[15] Marco Tulio Ribeiro, Sameer Singh, and Carlos Guestrin. ” why should i trust you?” explaining the predictions of any classifier. In Proceedings of the 22nd ACM SIGKDD international conference on knowledge discovery and data mining, pages 1135–1144, 2016.

[16] Scott M. Lundberg and Su-In Lee. A unified approach to interpreting model predictions. In Advances in Neural Information Processing Systems (NeurIPS), volume 30, pages 4765–4774, 2017.

[17] Julius Adebayo, Justin Gilmer, Michael Muelly, Ian Goodfellow, Moritz Hardt, and Been Kim. Sanity checks for saliency maps. Advances in neural information processing systems, 31, 2018.

[18] Mina Jamshidi Idaji, Julius Hense, Tom Neuhäuser, Augustin Krause, Yanqing Luo, Oliver Eberle, Thomas Schnake, Laure Ciernik, Farnoush Rezaei Jafari, Reza Vahidimajd, Jonas Dippel, Christoph Walz, Frederick Klauschen, Andreas Mock, and Klaus-Robert Müller. Beyond attention heatmaps: How to get better explanations for multiple instance learning models in histopathology. Medical Image Analysis, 113:104148, 2026.

[19] Cynthia Rudin. Stop explaining black box machine learning models for high stakes decisions and use interpretable models instead. Nature machine intelligence, 1(5):206–215, 2019.

[20] Bas H. M. van der Velden, Hugo J. Kuijf, Kenneth G. A. Gilhuijs, and Max A. Viergever. Explainable artificial intelligence (XAI) in deep learning-based medical image analysis. Medical Image Analysis, 79:102470, 2022.

[21] Pang Wei Koh, Thao Nguyen, Yew Siang Tang, Stephen Mussmann, Emma Pierson, Been Kim, and Percy Liang. Concept bottleneck models. In Proceedings of the 37th International Conference on Machine Learning (ICML), pages 5338–5348, 2020.

[22] Saarthak Kapse, Pushpak Pati, Srijan Das, Jingwei Zhang, Chao Chen, Maria Vakalopoulou, Joel Saltz, Dimitris Samaras, Rajarsi R. Gupta, and Prateek Prasanna. SI-MIL: Taming deep MIL for self-interpretability in gigapixel histopathology. In Proceedings of the IEEE/CVF Conference on Computer Vision and Pattern Recognition (CVPR), pages 11226–11237, 2024.

[23] Vitali Petsiuk, Abir Das, and Kate Saenko. RISE: Randomized input sampling for explanation of black-box models. In Proceedings of the British Machine Vision Conference (BMVC), 2018.

[24] Mukund Sundararajan, Ankur Taly, and Qiqi Yan. Axiomatic attribution for deep networks. In Proceedings of the 34th International Conference on Machine Learning (ICML), pages 3319–3328, 2017.

[25] Andrei Margeloiu, Matthew Ashman, Umang Bhatt, Yanzhi Chen, Mateja Jamnik, and Adrian Weller. Do concept bottleneck models learn as intended? arXiv preprint arXiv:2105.04289, 2021.

[26] Anita Mahinpei, Justin Clark, Isaac Lage, Finale Doshi-Velez, and Weiwei Pan. Promises and pitfalls of black-box concept learning models. arXiv preprint arXiv:2106.13314, 2021.

[27] Tuba N Gide, Camelia Quek, Alexander M Menzies, Annie T Tasker, Ping Shang, Jeff Holst, Jason Madore, Su Yin Lim, Rebecca Velickovic, Matthew Wongchenko, et al. Distinct immune cell populations define response to anti-pd-1 monotherapy and anti-pd-1/anti-ctla-4 combined therapy. Cancer cell, 35(2):238–255, 2019.

[28] Muhammad Shaban, Yuzhou Chang, Huaying Qiu, Yao Yu Yeo, Andrew H. Song, Guillaume Jaume, et al. A foundation model for spatial proteomics. arXiv preprint arXiv:2506.03373, 2025.

[29] Zongxin Yang, Linchao Zhu, Yu Wu, and Yi Yang. Gated channel transformation for visual recognition. In 2020 IEEE/CVF conference on computer vision and pattern recognition (CVPR), pages 11791–11800. IEEE, 2020.

[30] Maximilian Ilse, Jakub M. Tomczak, and Max Welling. Attention-based deep multiple instance learning. In Proceedings of the 35th International Conference on Machine Learning (ICML), pages 2127–2136, 2018.

