## Supplementary Tables and Figures for "CB-COMIL: interpretable prediction of melanoma immunotherapy response from multiplex immunofluorescence whole-slide images"

### Abstract

Deep learning models can predict melanoma immunotherapy response but rarely explain predictions biologically. Concept bottleneck models offer interpretable predictions, but have not been applied to multiplex immunofluorescence (mIF), and whether their learned concepts reproduce tissue measurements remains unclear. We developed CB-COMIL, which routes mIF predictions through four supervised concepts (immune:tumour ratio, CD8/SOX10 balance, CD68 relative signal and PD-L1 evasion index), decomposing each prediction exactly into concept contributions. Developed on 211 slides and evaluated on 42 slides from an independent, differently acquired test cohort, CB-COMIL achieved macro-F1 0.69 versus 0.67 for an opaque comparator (difference 0.020, 95% CI −0.103 to 0.144). In 41 slides with complete measurements, learned concepts correlated with measured counterparts (Spearman $\rho$ 0.61–0.76), yet substituting measured values reduced macro-F1 from 0.68 to 0.48. CB-COMIL makes predictions interpretable without detectable performance loss, and our evaluation shows that concepts can faithfully determine predictions without reproducing the corresponding biological measurements.

### Supplementary information

Table S1: Faithfulness of patch attributions under deletion and insertion tests on 42 slides. We compare CB-COMIL attention, integrated gradients (IG) and random patch rankings. Each entry is the area under the corresponding metric curve divided by the length of the evaluated fraction range (0 to 0.9 for deletion and 0 to 1 for insertion, main text Fig. 2). Bold indicates a more favourable value than the random control at the reported precision.

| | Deletion (↓) | | | Insertion (↑) | | |
|---|---|---|---|---|---|---|
| Metric | Attn | IG | Random | Attn | IG | Random |
| Macro-F1 | **0.56** | **0.45** | 0.68 | 0.68 | **0.69** | 0.68 |
| Recall (NR) | **0.51** | **0.53** | 0.89 | **0.89** | **0.89** | 0.88 |
| F1 (NR) | **0.50** | **0.46** | 0.72 | 0.72 | 0.72 | 0.72 |
| AUPRC | **0.64** | **0.63** | 0.77 | **0.78** | 0.77 | 0.77 |
| Flip rate[a] | **0.259** | **0.529** | 0.005 | **0.005** | **0.000** | 0.015 |

Lower values for deletion and higher values for insertion indicate more faithful rankings, except for flip rate. NR, non-responder. [a]Flip-rate is the proportion of predictions that changed under the perturbation; higher deletion and lower insertion flip-rates are favourable.

Table S2: Per-patient predictions and learned concept activations for the 42-WSI independent test cohort (anonymised IDs). Therapy indicates anti-PD-1 monotherapy (PD1) or combined anti-CTLA-4 and anti-PD-1 (IPI+PD1). True and Pred indicate observed and predicted outcomes (R, responder; NR, non-responder); P(R) denotes predicted response probability (threshold 0.5), and OK indicates correct classification. The four concepts are immune:tumour ratio, CD8/SOX10 balance, CD68 relative signal and PD-L1 evasion index. Concept values are learned activations on the scale of the biological targets (log1p-transformed and standardised using training-cohort statistics) and are the inputs to the prediction head and to the patient-level decompositions. They are not directly measured biomarker quantities. Several predictions lie close to the threshold (T07, T18, T24, 0.52; T33, 0.53); the most confident errors in each direction were T29, a non-responder predicted to respond ($P(\mathrm{R}) = 0.65$), and T19, a responder predicted not to respond ($P(\mathrm{R}) = 0.07$).

| ID | Therapy | True | Pred | P(R) | OK | imm:tum | CD8/SOX10 | CD68 rel. | PD-L1ev |
|---|---|---|---|---|---|---|---|---|---|
| T01 | IPI+PD1 | NR | NR | 0.38 | yes | -0.650 | 0.064 | -0.296 | -1.472 |
| T02 | PD1 | NR | NR | 0.08 | yes | -3.124 | -2.513 | 2.713 | -0.337 |
| T03 | IPI+PD1 | R | NR | 0.37 | no | -0.934 | -0.997 | 1.658 | 0.237 |
| T04 | IPI+PD1 | NR | NR | 0.33 | yes | -1.441 | -1.428 | 1.797 | 1.056 |
| T05 | PD1 | R | NR | 0.21 | no | -1.862 | -1.652 | 1.227 | 0.049 |
| T06 | IPI+PD1 | NR | NR | 0.09 | yes | -3.256 | -2.812 | 1.866 | 0.508 |
| T07 | IPI+PD1 | R | R | 0.52 | yes | 0.179 | 0.305 | -0.869 | -1.253 |
| T08 | PD1 | R | NR | 0.30 | no | -0.944 | -0.975 | 0.369 | -0.817 |
| T09 | PD1 | NR | NR | 0.36 | yes | -0.883 | -1.251 | 0.577 | 0.298 |
| T10 | PD1 | R | NR | 0.37 | no | -0.801 | -0.611 | 0.952 | -0.455 |
| T11 | IPI+PD1 | R | NR | 0.45 | no | -0.459 | -0.366 | -0.078 | -0.203 |
| T12 | PD1 | R | NR | 0.40 | no | -0.310 | -0.261 | 0.386 | -1.462 |
| T13 | IPI+PD1 | R | R | 0.58 | yes | 0.298 | 0.945 | -1.080 | -1.370 |
| T14 | PD1 | NR | NR | 0.42 | yes | -0.262 | -0.448 | 0.766 | -1.078 |
| T15 | PD1 | R | R | 0.64 | yes | 0.582 | 1.069 | -0.257 | -1.321 |
| T16 | IPI+PD1 | R | NR | 0.33 | no | -1.081 | -0.379 | 1.004 | -0.891 |
| T17 | PD1 | R | NR | 0.33 | no | -1.194 | -1.394 | 1.949 | 0.585 |
| T18 | IPI+PD1 | R | R | 0.52 | yes | -0.389 | -0.516 | 0.614 | 0.737 |

Table S2 continued

| ID | Therapy | True | Pred | P(R) | OK | imm:tum | CD8/SOX10 | CD68 rel. | PD-L1ev |
|---|---|---|---|---|---|---|---|---|---|
| T19 | PD1 | R | NR | 0.07 | no | -3.265 | -2.780 | 2.900 | -0.416 |
| T20 | IPI+PD1 | NR | NR | 0.36 | yes | -1.234 | -1.209 | 1.693 | 0.896 |
| T21 | IPI+PD1 | NR | NR | 0.24 | yes | -1.392 | -1.587 | 0.425 | -0.249 |
| T22 | PD1 | R | R | 0.57 | yes | 0.247 | -0.074 | 0.441 | -0.357 |
| T23 | PD1 | NR | NR | 0.40 | yes | -0.571 | -0.859 | 0.874 | -0.162 |
| T24 | IPI+PD1 | R | R | 0.52 | yes | -0.227 | -0.396 | 0.576 | 0.311 |
| T25 | IPI+PD1 | NR | NR | 0.19 | yes | -1.987 | -2.179 | 1.691 | 0.429 |
| T26 | PD1 | NR | NR | 0.41 | yes | -0.402 | 0.162 | -0.386 | -1.507 |
| T27 | PD1 | R | R | 0.54 | yes | 0.150 | 0.210 | 0.103 | -0.896 |
| T28 | IPI+PD1 | R | R | 0.59 | yes | 0.386 | 0.289 | -0.496 | -0.694 |
| T29 | IPI+PD1 | NR | R | 0.65 | no | 0.632 | 0.921 | -0.986 | -1.041 |
| T30 | IPI+PD1 | R | NR | 0.23 | no | -1.378 | -1.004 | 0.468 | -1.372 |
| T31 | IPI+PD1 | NR | NR | 0.46 | yes | -0.455 | -0.531 | 0.925 | -0.002 |
| T32 | PD1 | NR | NR | 0.43 | yes | -0.387 | -0.765 | 0.956 | -0.227 |
| T33 | IPI+PD1 | NR | R | 0.53 | no | -0.400 | -0.731 | 0.873 | 1.187 |
| T34 | PD1 | NR | NR | 0.32 | yes | -1.123 | -0.355 | 1.447 | -1.003 |
| T35 | IPI+PD1 | R | R | 0.82 | yes | 1.606 | 1.753 | -1.206 | -0.795 |
| T36 | IPI+PD1 | NR | NR | 0.20 | yes | -1.563 | -0.780 | 0.379 | -1.938 |
| T37 | IPI+PD1 | R | NR | 0.46 | no | -0.397 | 0.206 | 0.063 | -0.858 |
| T38 | PD1 | R | R | 0.60 | yes | 0.596 | 0.944 | -1.009 | -1.722 |
| T39 | IPI+PD1 | NR | NR | 0.18 | yes | -1.996 | -1.430 | 1.324 | -0.590 |
| T40 | IPI+PD1 | R | R | 0.55 | yes | 0.305 | 0.550 | -1.030 | -1.328 |
| T41 | IPI+PD1 | NR | NR | 0.35 | yes | -0.995 | -0.694 | 0.612 | -0.238 |
| T42 | PD1 | R | R | 0.70 | yes | 0.891 | 0.805 | -0.896 | -0.551 |

### Pairwise concept decision spaces

Supplementary Figure S3 shows the five concept pairs beyond the immune ratio and CD8 balance pair presented in the main text. The panels show how the predicted probability of response varies with each pair of learned concept values when the other two concepts are held fixed.

The supplementary panels show that the immune ratio and CD8 balance exert greater influence on predictions than the remaining two concepts. In panels involving relative CD68 signal, the decision boundary runs nearly parallel to the CD68 axis, so moving along that axis barely changes the prediction, consistent with its small coefficient in the prediction head. The boundary is more steeply inclined with respect to the PD-L1 evasion axis, consistent with this concept contributing to T33's borderline prediction. T29 falls on the response side of the boundary in every panel that includes the immune ratio or CD8 balance, consistent with these learned values driving its incorrect prediction of response.

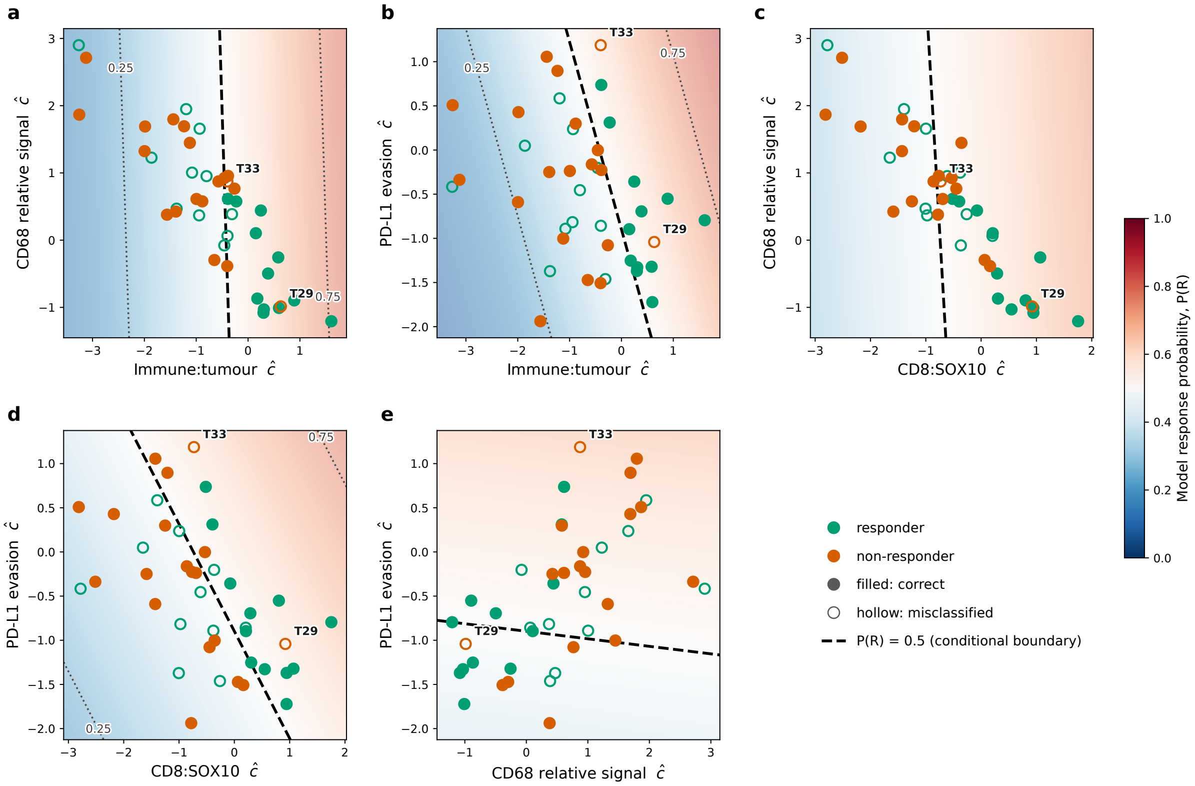


Figure S3: **Conditional decision spaces for the remaining pairs of learned concepts.** Each panel positions patients using their learned values for the two concepts shown on the axes. The background colour shows the response probability calculated with the other two concepts fixed at their cohort reference values. The dashed line marks the decision boundary at ($P(\mathrm{R}) = 0.5$). A patient's probability in a panel can differ from their full model prediction, which uses all four of that patient's learned concept values.

Table S4: **CB-COMIL compared with the opaque backbone, interpretable baselines on measured features, and fusion of measured features with the image embedding.** Opaque COMIL serves as a black box performance comparator without a concept bottleneck. Bold indicates the best value for each metric on the independent test cohort. Spec, specificity (non-responder recall); Sens, sensitivity (responder recall).

| | | **Cross-validation** | | | | **Independent test** | | | |
|---|---|---|---|---|---|---|---|---|---|
| **Model** | $n$ | **F1** | **Spec** | **Sens** | **AUPRC** | **F1** | **Spec** | **Sens** | **AUPRC** |
| **CB-COMIL** | 42 | 0.67 | 0.66 | 0.69 | 0.72 | **0.69** | **0.90** | 0.52 | **0.77** |
| Opaque COMIL | 42 | 0.67 | 0.62 | 0.74 | 0.71 | 0.67 | 0.68 | **0.65** | 0.70 |
| *Measured features only* | | | | | | | | | |
| Oracle (4 measured concepts)[a] | 41 | 0.60 | 0.56 | 0.63 | 0.70 | 0.54 | 0.89 | 0.30 | 0.70 |
| Interpretable panel, $L_1$[b] | 41 | 0.66 | 0.69 | 0.63 | 0.69 | 0.59 | 0.67 | 0.52 | 0.60 |
| *Measured features fused with the embedding* | | | | | | | | | |
| Intermediate fusion | 42 | 0.69 | 0.53 | 0.85 | 0.69 | 0.51 | 0.42 | 0.61 | 0.57 |
| Late fusion ($\alpha = 0.25$) | 42 | 0.64 | 0.62[c] | 0.74[c] | 0.67 | 0.59 | 0.58 | 0.61 | 0.66 |

Measured mIF features were unavailable for one patient in the independent test cohort, so comparisons requiring them are restricted to the 41 patients with a complete quantified panel. CB-COMIL achieved a macro-F1 of 0.68 on this subset. Predictions themselves require only the image and were available for all 42 patients.

[a] $L_2$-regularised logistic regression fitted on the measured values of the four predefined concepts.

[b] $L_1$-regularised logistic regression fitted on the full 21-feature handcrafted panel (main text Table 2), with regularisation strength tuned by internal cross-validation. This is the interpretable comparator.

[c] Late-fusion cross-validation specificity and sensitivity are inherited from the deep backbone's fold selection, as late fusion blends probabilities post hoc; only cross-validation macro-F1 was recomputed for the blend.

Table S5: CB-COMIL ablation results with uncertainty estimates for each model component, evaluated using five-fold cross-validation and the independent test cohort. Cross-validation values are reported as point estimates with SDs in superscript. Independent test cohort values are reported as point estimates with the upper and lower bounds of the 95% bootstrap CIs in superscript and subscript, respectively. F1, macro-F1; Spec, specificity (non-responder recall); Sens, sensitivity (responder recall); AUPRC.

| | Cross-validation | | | | Independent test | | | |
|---|---|---|---|---|---|---|---|---|
| Configuration | F1 | Spec | Sens | AUPRC | F1 | Spec | Sens | AUPRC |
| **CB-COMIL$^{\text{reference}}$** | $0.67^{\pm 0.03}$ | $0.66^{\pm 0.14}$ | $0.69^{\pm 0.16}$ | $0.72^{\pm 0.03}$ | $0.69^{0.85}_{0.57}$ | $0.90^{1.00}_{0.74}$ | $0.52^{0.73}_{0.32}$ | $0.77^{0.92}_{0.62}$ |
| *Encoder* | | | | | | | | |
| KRONOS$^{\text{global}}$ | $0.70^{\pm 0.06}$ | $0.60^{\pm 0.15}$ | $0.80^{\pm 0.08}$ | $0.72^{\pm 0.09}$ | $0.60^{0.74}_{0.44}$ | $0.68^{0.88}_{0.45}$ | $0.52^{0.72}_{0.32}$ | $0.75^{0.89}_{0.57}$ |
| ResNet-101 | $0.67^{\pm 0.05}$ | $0.67^{\pm 0.09}$ | $0.68^{\pm 0.14}$ | $0.73^{\pm 0.06}$ | $0.59^{0.74}_{0.44}$ | $0.58^{0.81}_{0.35}$ | $0.61^{0.80}_{0.40}$ | $0.58^{0.79}_{0.41}$ |
| DenseNet-121 | $0.62^{\pm 0.08}$ | $0.51^{\pm 0.12}$ | $0.73^{\pm 0.07}$ | $0.63^{\pm 0.06}$ | $0.49^{0.63}_{0.33}$ | $0.37^{0.60}_{0.16}$ | $0.61^{0.80}_{0.40}$ | $0.58^{0.81}_{0.41}$ |
| *Aggregator* | | | | | | | | |
| mean-pool | $0.66^{\pm 0.04}$ | $0.57^{\pm 0.13}$ | $0.81^{\pm 0.08}$ | $0.70^{\pm 0.11}$ | $0.57^{0.71}_{0.41}$ | $0.63^{0.84}_{0.41}$ | $0.52^{0.72}_{0.32}$ | $0.69^{0.85}_{0.52}$ |
| ABMIL$^{\text{flat}}$ | $0.67^{\pm 0.03}$ | $0.63^{\pm 0.15}$ | $0.74^{\pm 0.13}$ | $0.70^{\pm 0.07}$ | $0.38^{0.51}_{0.24}$ | $0.16^{0.35}_{0.00}$ | $0.65^{0.84}_{0.45}$ | $0.71^{0.86}_{0.53}$ |
| ABMIL$^{\text{per-marker}}$ | $0.67^{\pm 0.04}$ | $0.54^{\pm 0.09}$ | $0.80^{\pm 0.07}$ | $0.71^{\pm 0.04}$ | $0.56^{0.71}_{0.40}$ | $0.47^{0.71}_{0.25}$ | $0.65^{0.84}_{0.45}$ | $0.67^{0.85}_{0.48}$ |
| Bare linear head | $0.67^{\pm 0.06}$ | $0.66^{\pm 0.13}$ | $0.69^{\pm 0.13}$ | $0.73^{\pm 0.08}$ | $0.54^{0.69}_{0.38}$ | $0.47^{0.71}_{0.25}$ | $0.61^{0.81}_{0.40}$ | $0.64^{0.84}_{0.46}$ |
| Opaque COMIL | $0.67^{\pm 0.06}$ | $0.62^{\pm 0.13}$ | $0.74^{\pm 0.16}$ | $0.71^{\pm 0.05}$ | $0.67^{0.81}_{0.51}$ | $0.68^{0.88}_{0.47}$ | $0.65^{0.84}_{0.45}$ | $0.70^{0.88}_{0.52}$ |
| No instance loss | $0.67^{\pm 0.04}$ | $0.63^{\pm 0.13}$ | $0.72^{\pm 0.11}$ | $0.71^{\pm 0.03}$ | $0.60^{0.74}_{0.44}$ | $0.42^{0.65}_{0.20}$ | $0.78^{0.95}_{0.61}$ | $0.77^{0.92}_{0.60}$ |
| Per-class attention | $0.66^{\pm 0.07}$ | $0.63^{\pm 0.21}$ | $0.75^{\pm 0.21}$ | $0.72^{\pm 0.03}$ | $0.44^{0.59}_{0.30}$ | $1.00^{1.00}_{1.00}$ | $0.13^{0.28}_{0.00}$ | $0.77^{0.92}_{0.60}$ |

The reference CB-COMIL model comprises marker-aware KRONOS features, a COMIL aggregator and a concept bottleneck head, with class-agnostic attention and instance-loss supervision; each ablation modifies a single component while holding the others fixed. The encoder ablations substitute global KRONOS, ResNet-101 or DenseNet-121 features for marker-aware KRONOS; the aggregator ablations substitute mean-pooling, flat ABMIL or marker-aware (per-marker) ABMIL for COMIL; the loss ablation removes instance-level supervision; the attention ablation substitutes per-class for class-agnostic attention; and the head ablation (bare linear head) replaces the concept bottleneck with a linear classifier applied directly to the slide embedding. Opaque COMIL denotes the fully black-box comparator: the COMIL backbone with a non-linear multilayer-perceptron head and no concept bottleneck. Independent test cohort estimates used patient-level percentile bootstrap resampling ($B$ = 10,000; $n$ = 42).

Table S6: **Leave-one-concept-out ablation of CB-COMIL.** Each model was retrained after removing the indicated concept, using the same training configuration and five-fold development cross-validation. Macro-F1 values are reported as mean ± standard deviation (SD) across five folds. Changes in macro-F1 are calculated relative to the full four-concept model.

| **Concept removed** | **CV macro-F1 (mean ± SD)** | **Δ macro-F1** |
|---|---|---|
| None (all four concepts) | **0.668 ± 0.033** | – |
| Immune:tumour ratio | 0.656 ± 0.041 | −0.012 |
| CD8/SOX10 balance | 0.664 ± 0.043 | −0.004 |
| CD68 relative signal | 0.641 ± 0.053 | −0.027 |
| PD-L1 evasion index | 0.659 ± 0.040 | −0.009 |

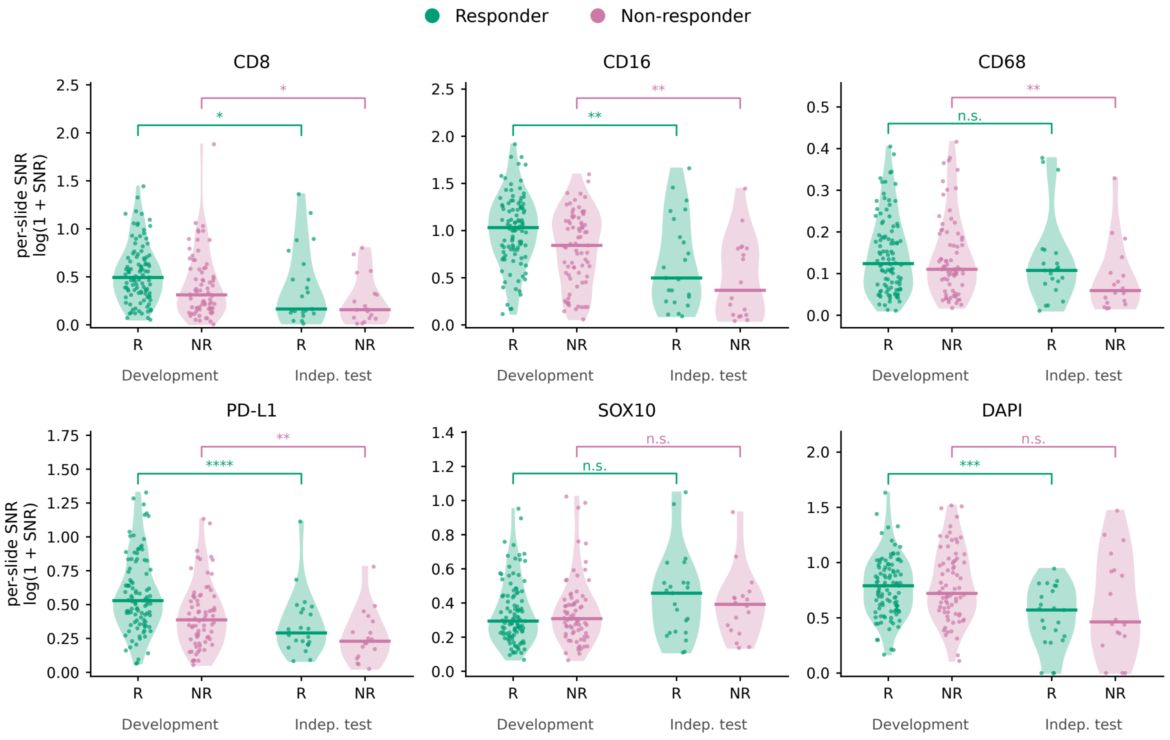


Figure S7: **Comparison of marker intensity distributions between development and independent test cohorts stratified by immunotherapy response.** Violin plots show the distributions of per-slide signal-to-noise ratios for six mIF channels (CD8, CD16, CD68, PD-L1, SOX10 and DAPI). Patients are grouped by cohort (development or test set) and treatment response (R, responder; NR, non-responder). Each point represents one patient, and horizontal lines indicate group medians. Brackets indicate comparisons between corresponding response groups across cohorts. Statistically significant differences were observed for CD8, CD16 and PD-L1 in both response groups, with additional differences for CD68 among non-responders and DAPI among responders. These findings demonstrate measurable differences in marker signal distributions between cohorts, including within response strata. n.s., not significant; *P ¡ 0.05, **P ¡ 0.01, ***P ¡ 0.001, ****P ¡ 0.0001.

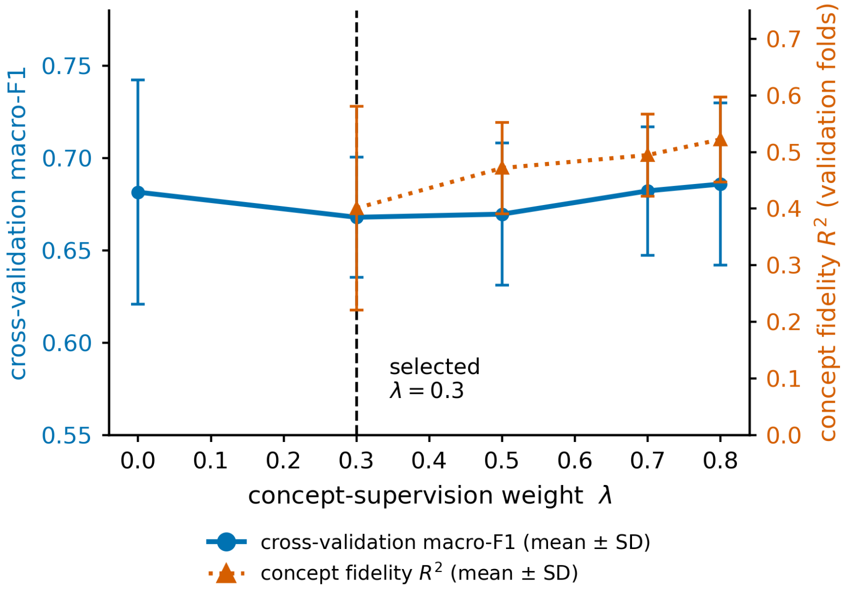


Figure S8: **Selection of the concept supervision weight $\lambda$.** Each point shows the mean across five validation folds, and error bars show one standard deviation across folds. The independent test cohort was excluded from this analysis. Concept fidelity (orange, right axis) is the mean $R^2$ across the four concepts. Higher $R^2$ indicates that the learned values more closely match the measured targets. Mean $R^2$ increased from 0.40 at $\lambda = 0.3$ to 0.52 at $\lambda = 0.8$, while macro-F1 for predicting immunotherapy response (blue, left axis) ranged from 0.668 to 0.686 across the nonzero weights. We selected $\lambda = 0.3$ to guide the learned concepts towards the measured targets while keeping the weight of the concept loss small. Larger weights improved concept fidelity, with little change in observed validation macro-F1.

Table S9: Robustness of CB-COMIL to random initialisation. Four training runs with different seeds, each using identical configuration with no per-seed tuning. Concept fidelity is the mean Spearman $\rho$ between learned activations and measured concepts across the 41 independent-test patients with complete mIF data. Macro-F1 is on the full independent test cohort ($n = 42$). Differences are CB-COMIL minus opaque COMIL, with 95% percentile confidence intervals from 10,000 patient-level bootstrap resamples. All results reported in the main text use seed 42.

| Seed | Fidelity $\rho$ | CB-COMIL | Opaque | $\Delta$ (95% CI) |
|---|---|---|---|---|
| 0 | 0.72 | 0.69 | 0.64 | +0.043 (−0.072, 0.163) |
| 1 | 0.71 | 0.63 | 0.60 | +0.031 (−0.118, 0.177) |
| 2 | 0.72 | 0.66 | 0.56 | +0.108 (−0.058, 0.278) |
| 42 | 0.69 | 0.69 | 0.67 | +0.020 (−0.103, 0.144) |
